# Inferring heterogeneous transmission and community introduction of antibiotic-resistant bacteria in hospital settings

**DOI:** 10.64898/2026.08.02.26359521

**Authors:** Jifan Li, Qing Yao, Sen Pei, Ning Ning

## Abstract

Antimicrobial-resistant organisms (AMROs) impose a major burden on healthcare systems, yet routine surveillance cannot readily distinguish colonization imported at admission from transmission acquired within hospitals. This gap is especially consequential because both processes may vary substantially across wards, while asymptomatic carriage, incomplete testing, imperfect diagnostic sensitivity, and patient movement obscure the underlying transmission dynamics. To address this challenge, we developed a blockwise agent-based iterated filter (BAIF) for inference in a patient-level transmission model on a dynamic ward co-location network. The model tracks susceptible and colonized patients as they move across wards, represents unobserved colonization histories, and incorporates the recorded testing schedule and imperfect diagnostic sensitivity. BAIF uses blockwise likelihood evaluation and resampling to estimate ward-block-specific transmission rates and importation probabilities in this high-dimensional latent system. Synthetic experiments showed that BAIF recovered these parameters from partially observed outbreaks. We then applied the framework to hospitalization and microbiological surveillance data collected from 2012 to 2016 at an urban quaternary care hospital in New York City for four AMROs. Transmission and importation were highly heterogeneous across ward blocks. Elevated transmission was repeatedly concentrated in the same ward groups, whereas blocks with the highest importation varied by pathogen. By distinguishing importation-dominated from transmission-dominated ward blocks, the framework can inform more targeted surveillance and infection-control strategies. More broadly, BAIF provides an effective inference framework for high-dimensional, partially observed agent-based models on dynamic contact networks.

**Significance Statement:** Antimicrobial-resistant organisms can enter hospitals with already-colonized patients or spread after admission, but routine testing often cannot tell these pathways apart. Many carriers have no symptoms, are never tested, or receive imperfect test results, and patients move among wards. We developed a new modeling approach that estimates heterogeneous, ward-group-specific hospital transmission and community introduction from patient movement and surveillance data. Applied to four resistant organisms at an urban quaternary care hospital in New York City, the method revealed large differences among ward groups in both imported colonization and within-hospital spread. Distinguishing these drivers can support more targeted control, including admission screening where importation is high and stronger infection-prevention measures where transmission is elevated.

## Introduction

Infections caused by antimicrobial-resistant organisms (AMROs) remain a major threat to patients and healthcare systems worldwide (1–4). Within hospitals, AMROs can spread through direct or indirect patient contact, contaminated equipment and surfaces, and interactions with healthcare workers (5–9). These dynamics are highly heterogeneous: organisms differ in their transmission characteristics, while both the risk of acquisition within the hospital and the likelihood of colonized patients entering from the community or other healthcare facilities vary across wards. Intensive care units (ICUs) illustrate this heterogeneity. Nosocomial infection rates in ICUs have been reported to be 5-10 times higher than those in general wards (10, 11), and ICU-based studies have documented cross-transmission of methicillin-resistant *Staphylococcus aureus* (12, 13), spread and persistence of vancomycin-resistant *Enterococcus faecalis* and *Enterococcus faecium* (14), and elevated transmission risk for other AMROs (15, 16). Such spatial variation can concentrate colonization pressure in particular wards or ward groups, creating localized hotspots where targeted interventions may have the greatest impact. Quantifying ward-level differences in transmission and importation is therefore essential for designing efficient and effective infection-control strategies.

Routine surveillance rarely captures this complexity. Only a fraction of patients are tested because many individuals carry AMROs asymptomatically and therefore escape routine detection. Diagnostic sensitivity may also be imperfect, while frequent transfers between wards continually reshape patient contact patterns. Consequently, the true burden of colonization and the relative contributions of importation and onward transmission remain poorly characterized (17–19). Simple indicators, such as length of stay or contact counts, and aggregate models that ignore heterogeneous, time-varying ward structure cannot reliably disentangle organism- and ward-specific transmission from importation using sparse and noisy surveillance data.

A substantial body of work has used mathematical models to characterize AMRO transmission and evaluate infection-control strategies in healthcare settings (5, 20–23). Under partial observation, several approaches have estimated key transmission processes, including nosocomial transmissibility and importation from the community (17, 19, 24). More recently, patient-movement networks and individual-level data have enabled analyses at finer spatial and temporal resolution (8, 25–33). Despite these advances, inference at the patient level remains challenging because colonization histories are largely unobserved, creating a high-dimensional latent process. Allowing transmission and importation to vary across ward blocks further increases the computational challenge.

To address these challenges, we develop an inference framework that integrates individual-based transmission modeling with an explicit representation of the surveillance process. Patients are modeled as susceptible or colonized on a dynamic ward co-location network reconstructed from hospitalization records, while the observation model accounts for incomplete testing and imperfect diagnostic sensitivity. Inference is performed using a blockwise agent-based iterated filter (BAIF), an iterated block-particle-filtering approach designed for agent-based transmission models on dynamic contact networks. By organizing likelihood evaluation and resampling at the ward-block level, BAIF enables high-dimensional inference under partial observation and jointly estimates block-specific transmission rates and importation probabilities while preserving patient-level colonization trajectories (34–39). We first evaluate parameter recovery through simulation studies under known transmission and importation scenarios. We then apply BAIF to hospitalization and microbiological surveillance data collected from 2012 to 2016 at an urban quaternary care hospital in New York City, focusing on four clinically important AMROs: methicillin-resistant *Staphylococcus aureus* (MRSA), carbapenem-resistant *Klebsiella pneumoniae* (CRKP), vancomycin-resistant *Enterococcus faecalis* and *Enterococcus faecium* (VRE), and levofloxacin-resistant *Pseudomonas aeruginosa* (LRPA). These organisms are prominent in infection-control guidance and global estimates of antimicrobial-resistance burden and were among the most frequently detected resistant organisms in our data (3, 4, 40, 41). By partitioning wards into communities defined by the patient-transfer network, we estimate spatially heterogeneous transmission and importation parameters. We identify pronounced heterogeneity: certain ward blocks serve as disproportionate reservoirs, and different organisms exhibit distinct spatial signatures. Our results establish a generalizable framework for inference in partially observed, high-dimensional agent-based models and provide epidemiological insights essential for designing efficient, targeted interventions against antimicrobial resistance in hospital settings.

## Results

### Observed Detection Patterns of AMROs

We first characterized observed detections of MRSA, CRKP, VRE, and LRPA from 2012 to 2016. Across all four organisms, weekly detections were sparse relative to the hospital population and varied substantially over time. Because surveillance was conducted through routine clinical testing, only a subset of patients was observed, and imperfect diagnostic sensitivity further limited detection of colonized patients. Panels A and B of Fig. 1 illustrate these constraints for MRSA: testing coverage varied markedly across wards and weeks, and positive detections were correspondingly sparse and unevenly distributed. Thus, the observed data provide only a partial and heterogeneous view of the underlying colonization process. Additional ward- and block-level surveillance summaries are provided in SI Appendix, Hospitalization and Surveillance Data (Figs. S1 and S2).

**Figure 1.**
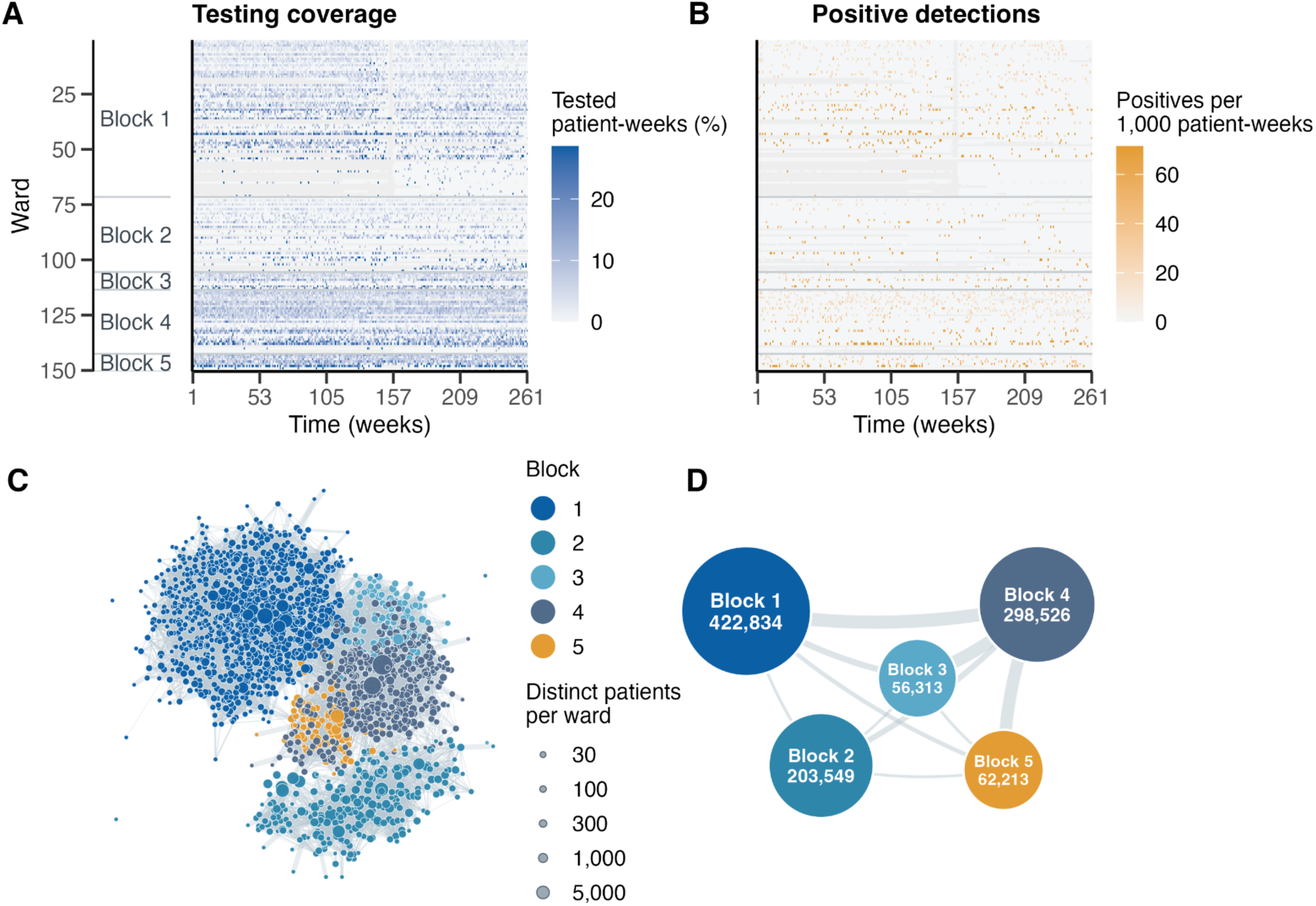
Ward-level MRSA surveillance and movement-based partition of the ward transfer network, 2012–2016. **(A)** Weekly testing coverage, calculated as the percentage of patient-weeks with an MRSA test among all patient-weeks present in each ward. **(B)** Weekly MRSA-positive detections per 1,000 patient-weeks. Wards are grouped and ordered by ward block. Only wards with at least 500 patient-weeks during the study period are shown. Gray cells indicate ward-weeks with no patients present. **(C)** Ward-level transfer network reconstructed from hospitalization records. Nodes represent wards, node size indicates the number of distinct patients who occupied each ward during 2012-2016, and edges represent patient transfers, with width proportional to normalized transfer-network weight. Node colors indicate ward-block membership identified by the Infomap algorithm. **(D)** Block-level network obtained by aggregating wards into five movement-based communities. Node size and labels indicate the number of patient-ward stays in each block, and edges represent transfers between blocks, with width proportional to aggregate normalized transfer-network weight. This partition defines the spatial scale for estimating block-specific transmission and importation parameters.

**Figure 2.**
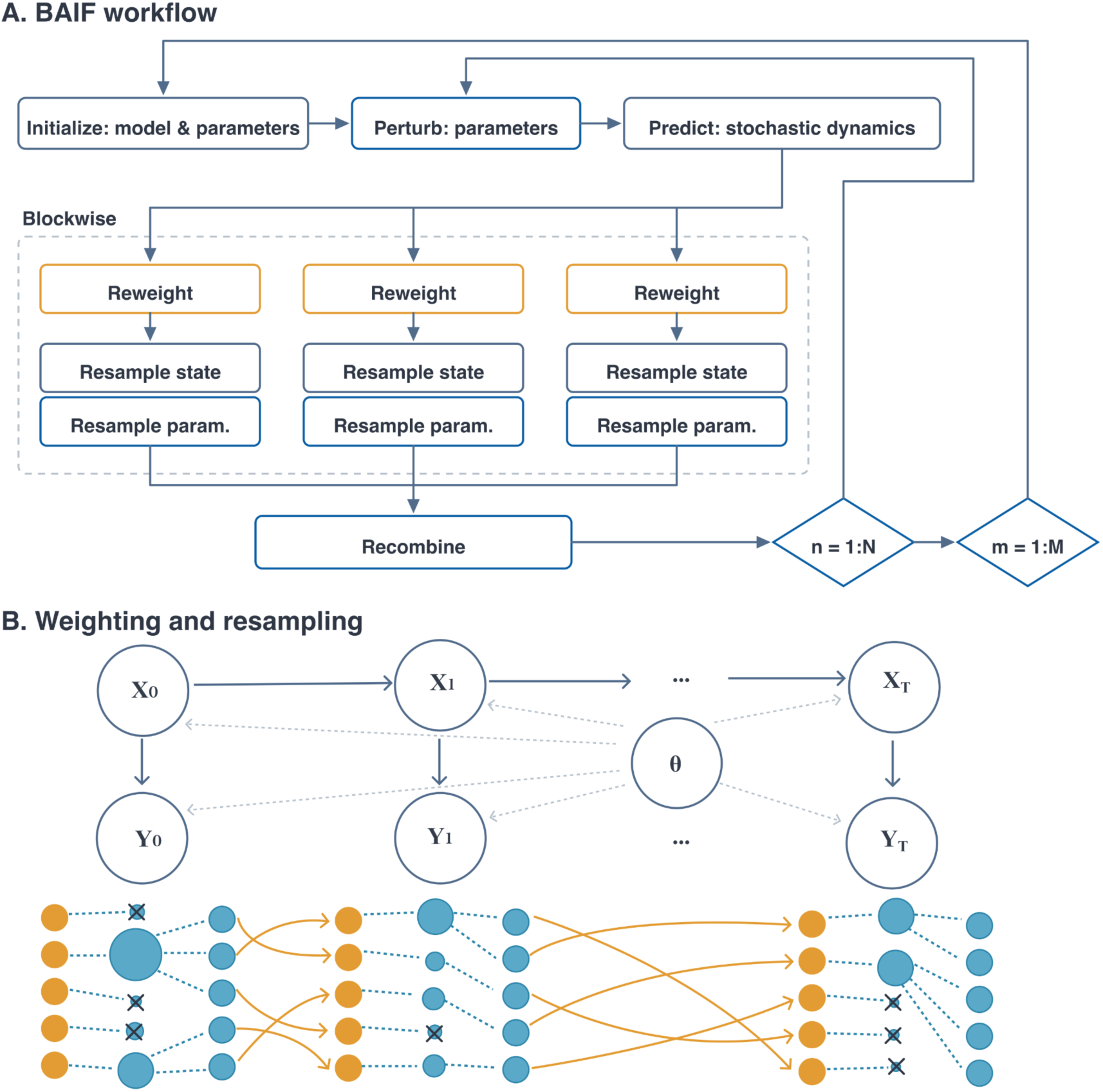
Overview of the blockwise agent-based iterated filter (BAIF). **(A)** BAIF workflow for particle propagation and parameter updating. Here, *n* indexes observation times and *m* indexes filtering iterations. Each particle carries a candidate realization of patient-level colonization states together with block-specific parameters, including the transmission rate *β*_g_ and importation probability *C*_0,g_. Particles are propagated through the agent-based transmission model on the dynamic ward co-location network, weighted within blocks, resampled, and recombined before the next update. **(B)** Blockwise weighting and resampling. *X_t_* denotes latent colonization states at time *t*, *Y_t_* denotes the corresponding observations, and *θ* denotes the parameter vector. Simulated and observed detections are compared separately within each ward block, generating block-specific weights used to update the corresponding latent states and parameters.

At the ward level, both testing intensity and positive detections varied widely across space and time. However, descriptive measures such as observed counts, patient-days, or crude detection rates cannot determine whether elevated burden in a ward reflects greater importation at admission, increased onward transmission, or more intensive testing. This ambiguity is compounded by asymptomatic carriage and incomplete observation. Consequently, raw surveillance patterns alone are insufficient to identify transmission hotspots or disentangle community introduction from within-hospital spread, motivating a mechanistic inference framework that explicitly accounts for patient movement, testing, and unobserved colonization.

### Ward Transfer Network and Block Definition

We reconstructed a ward-transfer network from hospitalization records and applied the Infomap community detection algorithm (42) to partition wards into five movement-based blocks for inference. Rather than estimating separate transmission and importation parameters for each ward, we grouped wards according to patient-transfer connectivity, thereby reducing the parameter dimension while preserving major patterns of patient flow. This movement-based grouping is epidemiologically relevant because wards connected by frequent transfers share patient-flow pathways and generate overlapping opportunities for co-location and transmission.

The five blocks contained 1,032, 256, 112, 434, and 100 wards, respectively. From 2012 to 2016, they accounted for 422,834 (40.5%), 203,549 (19.5%), 56,313 (5.4%), 298,526 (28.6%), and 62,213 (6.0%) patient-ward stays, respectively (Fig. 1 C and D). Overall, 84.2% of ward-to-ward transfers occurred between wards assigned to the same block, indicating that the partition captured the dominant patient-flow structure while substantially reducing the spatial dimension of inference. Network construction and patient-block assignment are detailed in SI Appendix, Ward Transfer Network and Block Construction (Fig. S3 and Tables S2 and S3).

Transmission and importation parameters were defined at the block level, such that all patients assigned to a block shared the same transmission rate and importation probability. Each patient was assigned to the block containing the greatest number of their distinct recorded patient-days. Subsequent transfers altered the set of contemporaneously co-located patients and therefore the transmission opportunities represented in the dynamic contact network.

### Agent-Based Modeling of Transmission and Importation

The ward-block partition defines the spatial scale for transmission and importation parameters, while patients’ time-varying ward locations determine the underlying contact structure. We modeled colonization using an individual-level agent-based framework on a dynamic ward co-location network reconstructed from hospitalization records. Patients are connected when they occupy the same ward at the same time, representing opportunities for direct contacts and indirect transmission through shared healthcare workers, equipment, and ward environments.

Each patient is classified as susceptible or colonized. Susceptible patients may acquire colonization through co-location with colonized patients in the same ward, with transmission governed by the block-specific parameter *β*_g_ for the ward block *g*. Colonized patients may subsequently clear colonization. Patients may also enter the hospital already colonized, represented by the block-specific importation probability *C*_0,g_. Observations arise through routine surveillance, in which only a subset of patient-weeks is tested and diagnostic sensitivity is imperfect. Details on the observation model are provided in Materials and Methods and SI Appendix, Agent-Based Colonization Model. This formulation separates colonization present at admission from colonization acquired through within-hospital transmission (18, 19, 24).

To infer the latent colonization process and estimate *β*_g_ and *C*_0,g_, we developed a blockwise agent-based iterated filter (BAIF). BAIF represents uncertainty with an ensemble of particles, each encoding a candidate reconstruction of patient-level colonization histories together with the associated block-specific transmission and importation parameters. For each particle, the agent-based model is propagated forward on the dynamic ward co-location network to generate predicted detections for comparison with the surveillance data. During filtering, simulated and observed block-level cases are compared separately within each ward block, yielding block-specific particle weights based on how well each particle reproduces the observations in that block. These weights are then used to update the corresponding latent states and block-specific parameters, including *β*_g_ and *C*_0,g_. Repeated cycles of propagation, weighting, resampling, and parameter perturbation progressively concentrate the ensemble in regions of parameter space that are consistent with the observed data. Detailed algorithmic and implementation specifications are provided in Materials and Methods and SI Appendix, Blockwise Agent-Based Iterated Filtering (Algorithm S1 and Table S4).

### Validation of Inference Using Simulated Data

We first evaluated BAIF using synthetic outbreaks generated under known block-specific transmission rates and importation probabilities. We then applied BAIF to the partially observed simulated data and assessed whether the estimated parameter distributions converged toward the true values across filtering iterations. This analysis tested the algorithm’s ability to recover block-specific transmission and importation parameters despite incomplete observation of the underlying colonization process. We repeated the validation across multiple combinations of *β*_g_ and *C*_0,g_ to evaluate robustness under a range of transmission and importation scenarios.

Figure 3 summarizes parameter recovery across four prespecified combination of block-specific *β*_g_ and *C*_0,g_ values. Panel A shows convergence for settings A–C separately for each of the five ward blocks. At early iterations, the parameter distributions are widely dispersed, reflecting the broad initial parameter ensemble. As filtering proceeds, the distributions contract toward the horizontal reference line at one, indicating exact recovery. Importation probabilities, *C*_0,g_, stabilize within fewer iterations, whereas transmission rates, *β*_g_, converge more gradually. This difference is consistent with importation being informed directly at admission, while transmission must be inferred from partially observed within-hospital colonization dynamics. By the final iterations, both parameter sets are centered near their true values across settings and blocks. The intervals contained the true value for 16 of 20 transmission-rate values and 19 of 20 importation-probability values. A detailed block-specific convergence example for setting D and the joint recovery of both parameters are provided in the SI Appendix, Synthetic Validation Design (Table S5 and Figs. S4 and S5).

**Figure 3.**
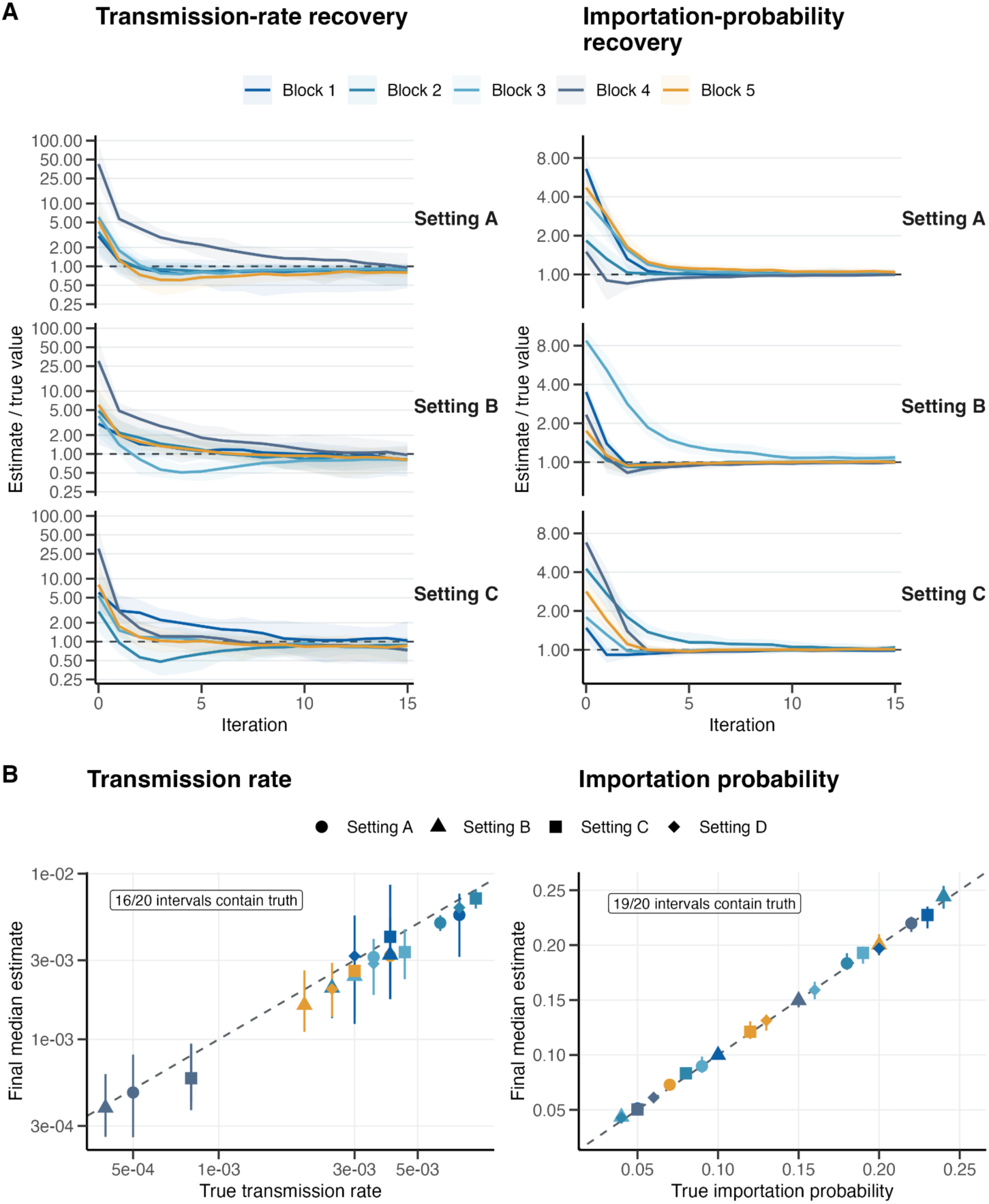
Recovery of block-specific parameters from synthetic outbreaks using BAIF. **(A)** The left panel shows recovery of transmission rates, *β*_g_, and the right panel shows recovery of importation probabilities, *C*_0,g_, for settings A–C. Lines show medians, and shaded bands denote the 2.5th–97.5th percentile intervals across 20 independent BAIF inference realizations per setting. Each trajectory is divided by its setting- and block-specific true value; the horizontal dashed line at one denotes exact recovery, and iteration 0 shows the initial parameter distribution before BAIF updating. **(B)** The left panel shows final transmission-rate estimates and the right panel shows final importation-probability estimates plotted against the true values across settings A–D and all five blocks. Points indicate medians, and vertical lines indicate the 2.5th–97.5th percentile intervals across 20 independent BAIF inference realizations per setting. Dashed identity lines denote equality between the final estimate and true value. Colors denote ward blocks, and shapes denote parameter settings. The true value lies within 16 of 20 transmission-rate intervals and 19 of 20 importation-probability intervals.

### Block-Specific Transmission and Importation Parameters for AMROs

After validating parameter recovery in synthetic outbreaks, we applied BAIF to the observed AMRO data to estimate block-specific transmission rates and importation probabilities in real-world hospital settings. For each pathogen, inference was conducted sequentially for each year from 2012 through 2016. Within each inference chain, the terminal latent colonization state from one year was carried forward to initialize the following year, preserving continuity in the underlying colonization process across the study period.

Figure 4A summarizes the distributions of five-year mean block-specific transmission and importation estimates across inference chains. The spatial pattern of transmission varied markedly by pathogen. Block 1 showed the highest transmission rate for all four pathogens, with relatively high rates also observed for MRSA in blocks 2 and 3, LRPA in blocks 2 and 5, and VRE in blocks 4 and 5. Importation probabilities followed a more consistent pattern across pathogens: blocks 1 and 4 generally had higher estimates, block 2 tended to have lower estimates, and block 3 showed greater variability. Overall, both transmission and importation were strongly heterogeneous across ward blocks, but the highest-risk blocks differed by pathogen and process. These findings suggest that infection-control priorities should be tailored to both pathogen and mechanism, with admission- focused screening in blocks with elevated importation and intensified transmission-control measures in blocks with greater within-hospital spread. Numerical summaries and diagnostics across inference chains are provided in SI Appendix, Summaries of Real-Data Parameter Estimates (Table S6 and Figs. S6 and S7).

**Figure 4.**
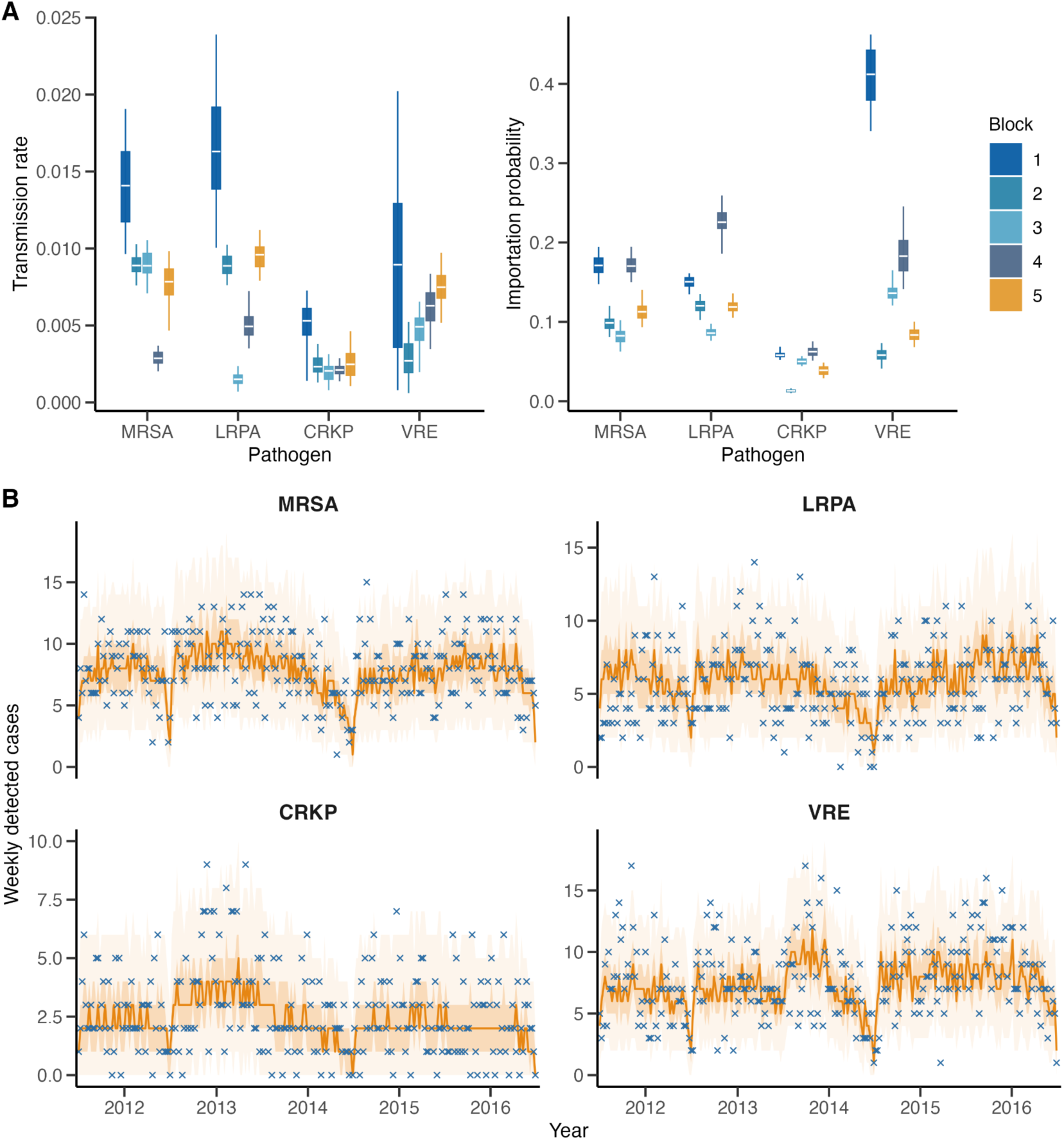
Block-specific parameter estimates and fitted-model simulations reproduce observed weekly AMRO detections, 2012–2016. **(A)** The left panel shows transmission rates, *β*_g_, and the right panel shows importation probabilities, *C*_0,g_, for each pathogen and ward block. For each inference chain, pathogen, and block, yearly estimates from 2012 to 2016 were averaged to obtain a five-year mean estimate. Colored box-and-whisker plots summarize the distribution of these chain-specific five-year means. Boxes span the interquartile range, center lines indicate medians, and whiskers extend from the 2.5th to the 97.5th percentiles, obtained from 100 independent inference realizations. Colors denote ward blocks. **(B)** Blue crosses indicate observed weekly detection counts, and orange lines show the median simulated detections generated from the fitted model under the recorded testing schedule. Dark and light shaded bands denote the central 50% and 95% simulation intervals, respectively. For each of the 100 fitted inference chains, 500 independent Poisson observation draws were generated from each weekly expected detection count. Panels correspond to MRSA, LRPA, CRKP, and VRE.

### Simulations Reproduce Observed AMRO Detections

To evaluate whether the fitted model reproduced the observed detection patterns, we generated ensembles of simulated detection trajectories using the fitted yearly parameters from each inference chain and the recorded testing schedules. Figure 4B shows that the fitted simulations broadly reproduced the magnitude and variability of weekly detections across pathogens. CRKP detections remained consistently sparse, MRSA and LRPA showed intermediate sustained levels, and VRE exhibited the highest burden with several periods of increased activity. The fitted median trajectories captured broad changes in detection levels while smoothing short-term fluctuations. From these simulations, we constructed week-specific 95% simulation intervals and assessed their empirical coverage of the observed data. The intervals contained 98.9% of weekly detections for MRSA, 97.3% for LRPA, 98.9% for CRKP, and 96.2% for VRE. Overall, the model reproduced detection patterns across pathogens with substantially different burdens and temporal variability. Simulation construction and coverage are detailed in SI Appendix, Fitted-Model Simulations and Contribution Decomposition (Table S7).

### Estimating the Contributions of Importations and Transmission

Because the fitted model reproduces the aggregate weekly detections for each pathogen, we used it to decompose colonization burden into two underlying sources: importation at admission and onward within-ward transmission. This distinction is operationally important because importation- dominated blocks may benefit most from admission screening or early testing, whereas transmission-dominated blocks may require intensified infection-control measures. The block-specific parameters *β*_g_ and *C*_0,g_ cannot be compared directly because they operate on different scales. We therefore quantified their model-estimated contributions to colonization burden using the fitted-model simulation ensemble. For each block and pathogen, contributions were summarized as the mean number of colonized patients per 1,000 patients present in the hospital over 2012 to 2016.

Figure 5 compares the estimated contributions of importation and within-hospital transmission within each ward block. Colonization burden was unevenly distributed across blocks for all four pathogens. Block 1 accounted for the largest share of the five-year burden in every pathogen, ranging from 41% for LRPA to 68% for VRE. Block 4 was consistently the second-largest contributor, accounting for 21%–34%. The remaining blocks contributed substantially less, although their relative importance varied by pathogen: block 2 contributed more to MRSA and LRPA than to CRKP or VRE, whereas blocks 3 and 5 remained comparatively small contributors across all pathogens. The corresponding block-specific values are provided in SI Appendix, Table S8.

**Figure 5.**
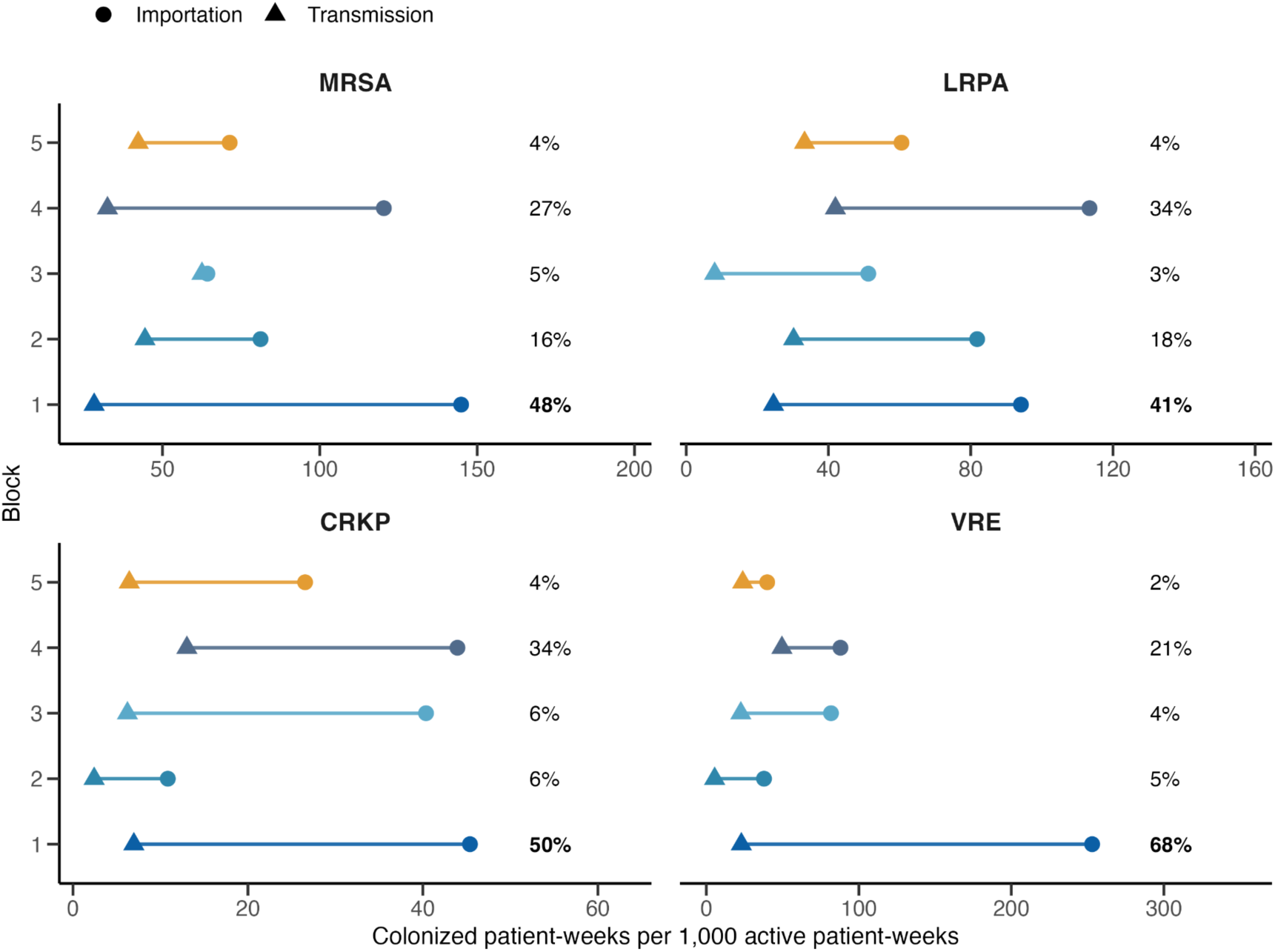
Block-specific contributions of importation and within-hospital transmission to colonization burden. For each pathogen and ward block, circles indicate the mean weekly colonization burden attributable to importation at admission, and triangles indicate the burden attributable to onward transmission. Values are averaged over 2012–2016 and reported as colonized patient-weeks per 1,000 active patient-weeks in each block. The percentage shown at the right of each row denotes that block’s share of the pathogen-specific total colonization burden over the study period; percentages sum to 100% within each pathogen panel.

Importation contributed more than transmission in every pathogen-block combination, but the relative balance varied across blocks and organisms. In block 1, VRE showed substantially greater importation dominance than the other three pathogens: its importation contribution far exceeded its transmission contribution and either contribution in every other VRE block. In block 3, importation and transmission were nearly equal for MRSA, whereas importation remained substantially larger for LRPA, CRKP, and VRE, indicating an unusually large relative contribution of within-hospital transmission to MRSA burden in this block. These patterns reveal substantial heterogeneity in both the spatial distribution of colonization burden and the relative contributions of importation and within-hospital transmission.

## Discussion

Our analysis reveals substantial heterogeneity in both within-hospital transmission and importation at admission across ward blocks. Across pathogens, fitted transmission rates varied several-fold among blocks, while median importation probabilities ranged from approximately 1% to 41%. These differences persisted after accounting for asymptomatic colonization, incomplete testing, imperfect diagnostic sensitivity, and time-varying patient movement. Colonization pressure was therefore unevenly distributed across the hospital system, and the balance between importation and onward transmission differed by block and pathogen. Such distinctions cannot be recovered reliably from raw detections alone, because routine surveillance combines multiple underlying processes into the same observed case counts (17–19).

Separating importation from onward transmission has direct implications for control. Ward blocks with a high burden driven largely by colonized patients entering the hospital may benefit most from admission screening, earlier testing, and rapid implementation of precautionary measures. By contrast, blocks in which within-hospital spread contributes more substantially may warrant intensified efforts to interrupt transmission, including strengthened isolation practices (40, 41), enhanced environmental cleaning (41, 43, 44), and targeted surveillance (45, 46). By allowing both processes to vary across blocks, our framework provides a more mechanistic explanation for why colonization burden is concentrated in particular parts of the hospital system and helps align interventions with the processes most responsible for that burden.

The cross-pathogen comparison further suggests that hospital risk includes both shared spatial structure and pathogen-specific dynamics. Blocks 1 and 4 consistently showed elevated colonization burden across organisms even after standardization by block size, indicating that these parts of the hospital network may represent recurring areas of elevated risk. However, similar burdens can arise through different mechanisms. A large transmission contribution may reflect either a high transmission rate or a high level of importation that creates more opportunities for onward spread; the fitted parameters are therefore needed to interpret the contribution estimates. The observed contrasts are also biologically plausible. The broader importation patterns for MRSA and VRE may reflect their established reservoirs in healthcare and long-term-care settings (47), whereas patterns for LRPA may be shaped by prior antibiotic exposure, healthcare contact, and environmental persistence in long-stay or device-exposed populations (48, 49). CRKP showed lower importation probabilities, consistent with a more concentrated acute-care-associated ecology (50). Thus, ward blocks that are repeatedly prominent across pathogens may still require organism-specific surveillance and control strategies.

The primary methodological contribution is BAIF, a likelihood-based framework for inference in partially observed agent-based transmission models with patient turnover and dynamic contact networks. The method is designed for settings in which patients are admitted, discharged, and transferred over time, colonization states are only sparsely observed, and transmission and importation must be estimated jointly without aggregating the latent patient-level process. In this setting, the state dimension grows with the number of patients, making standard global particle-filtering approaches vulnerable to weight degeneracy and the curse of dimensionality. BAIF addresses this challenge by using ward-block structure to organize parameter sharing, likelihood evaluation, and resampling, while retaining patient-level colonization states throughout inference. Rather than applying a single global correction across the entire hospital network, BAIF uses local block-specific likelihood information to guide state updating and parameter learning. This construction enables joint estimation of block-specific transmission and importation parameters in a high-dimensional hospital model with dynamic contacts, patient turnover, and incomplete surveillance.

Several limitations should be considered. Surveillance was based on routine clinical care rather than systematic screening, so inference was constrained by which patients were tested, when testing occurred, and the sensitivity of the diagnostic process. The transmission model represented ward co-location as the primary exposure mechanism and did not explicitly distinguish transmission mediated by healthcare workers, environmental reservoirs, or other organism-specific pathways (6, 7, 9). In addition, the analysis focused on inference and interpretation rather than formal optimization of intervention strategies. Privacy restrictions also prevented us from linking de-identified ward blocks to specific clinical settings, such as emergency departments and intensive care units, limiting clinical interpretation of block-level differences.

These limitations point to several directions for future work. The framework could be used to assess whether high-risk blocks remain stable over time and to compare the effects of targeting admission screening, intensified surveillance, or transmission-reduction measures to different parts of the hospital network. It also provides a foundation for prospective simulation studies that evaluate block-specific intervention strategies under incomplete surveillance (51–53). More broadly, BAIF offers an inference framework for characterizing heterogeneous hospital transmission and for informing the design of targeted infection-control strategies.

## Materials and Methods

### Data and surveillance process

We analyzed hospitalization records and microbiological surveillance data collected as part of clinical care from a New York hospital system between 2012 and 2016. Hospitalization records include patient admissions, discharges, and ward locations, which were used to reconstruct patient movement and co-location patterns over time. Microbiological data record detections of four antimicrobial-resistant organisms: methicillin-resistant *Staphylococcus aureus* (MRSA), levofloxacin-resistant *Pseudomonas aeruginosa* (LRPA), carbapenem-resistant *Klebsiella pneumoniae* (CRKP), and vancomycin-resistant *Enterococcus faecalis* and *Enterococcus faecium* (VRE). Surveillance was conducted as part of routine clinical practice. Only a subset of patient-weeks was tested, and diagnostic sensitivity was imperfect. Consequently, observed detections represent a partial and noisy observation of underlying colonization dynamics. These testing schedules and sensitivities were explicitly incorporated into the observation model described below.

### Dynamic contact network

We represented patient contact structure using a dynamic ward co-location network reconstructed from hospitalization records. In this network, nodes represent individual patients, and undirected edges connect pairs of patients occupying the same ward contemporaneously. Such co-location captures opportunities for direct transmission and indirect transmission mediated by shared healthcare workers and ward environments, without requiring explicit observation of individual contacts (6, 7, 28). The network evolves over time as patients are admitted, discharged, and transferred between wards. Colonization dynamics are simulated on this time-varying network through seven daily substeps per week.

### Colonization transmission model

We modeled colonization dynamics using an individual-level agent-based framework. Each patient occupies one of two states: susceptible (*S*) or colonized (*C*). Within the hospital, susceptible patients may acquire colonization through co-location with colonized patients in the same ward. Transmission is governed by a block-specific transmission parameter *β*_g_, reflecting heterogeneity in transmission intensity across ward groups. Colonized patients may clear colonization spontaneously at rate *α*. In addition, patients may enter the hospital already colonized, representing importation from the community or other healthcare settings. This process is modeled via a block-specific importation probability *C*_0,g_ applied at admission.

Let *g*(*i*) denote the block assigned to patient *i*, *N_i_*(*t*) denote the set of contemporaneous ward co-location neighbors of patient *i*, and *P*{*C_i_*(*t*) = *1*} denote the colonization probability of patient *i* at time *t*. For the admission time *t_adm_*, the admission model is

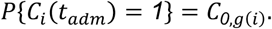

After admission, the acquisition hazard for a susceptible patient is determined by the corresponding block-specific transmission rate and the number of colonized neighbors,

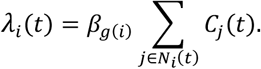

Within each daily step, the colonization probability is updated according to

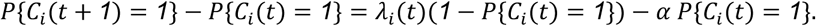

Here *α* is the spontaneous clearance parameter for a daily step. We held *α* fixed during inference, using *α* = *0*.*003* per day for MRSA and CRKP, *α* = *0*.*016* per day for LRPA, and *α* = *0*.*017* per day for VRE. Under this discrete-time clearance step, these values imply median durations of colonization or detectable carriage *log*(*0*.*5*)/*log*(*1* − *α*) of approximately 231, 231, 43, and 40 days, respectively, chosen to fall within published ranges reported for these organisms or related resistant phenotypes (18, 54–57).

### Ward grouping into blocks

We used patient-transfer connectivity to define the spatial scale because transfers are the observed movement pathways by which colonized patients can enter new wards and subsequently be co-located with other patients. We constructed a ward-to-ward transfer network in which nodes represent wards and edge weights correspond to normalized patient transfer frequencies between wards (25, 26, 30, 53).

Specifically, for each ordered pair of wards (*u* → *v*), we computed

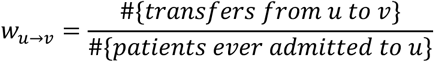

and defined the undirected edge weight as *w_uv_* = *w_u_*_→*v*_ + *w_v_*_→*u*_. Community detection was performed on this weighted network using the Infomap algorithm (42) to define ward blocks for inference. The partition contained 1,032, 256, 112, 434, and 100 wards across the five blocks, respectively, with 84.2% of ward-to-ward transfers occurring within blocks. Each patient was assigned to the block containing the greatest number of that patient’s distinct recorded patient-days. Transmission and importation parameters were then defined at the block level.

### Observation model

Observed detections arise through routine surveillance. At each patient-week, testing occurs according to the recorded testing schedule. Let *T_i_*(*t*) indicate whether patient *i* was tested at time *t*, and *Y_i_*(*t*) denote the observed detection status. The observation model is

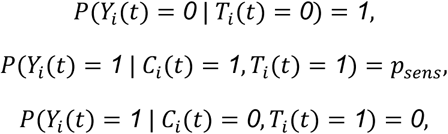

where *p_sens_* = 0.4 is diagnostic sensitivity and the test specificity is assumed to be 1 (18). This observation model links latent colonization states to observed detections and is applied consistently in both inference and simulation.

### Inference via the blockwise agent-based iterated filter

We inferred block-specific transmission and importation parameters using the blockwise agent-based iterated filter (BAIF). The overall BAIF workflow is summarized in Fig. 2. BAIF adapts earlier iterated block-filtering ideas to the hospital setting studied here (34, 35, 39, 58). Unlike earlier aggregated particle-filtering applications, each particle in BAIF carries patient-level colonization states on a weekly updated contact network, with admissions, discharges, and transfers changing the active contact structure over time. Within each filtering step, a likelihood is computed for each block and used to update that block’s importation and transmission parameters under partial observation. At each iteration, the parameters are perturbed using a decreasing-variance random walk, and the resulting ensemble of state-parameter particles is then propagated forward according to the transmission model. Likelihoods are computed by comparing simulated detections with observed data under the observation model, evaluated independently within each block. Particles are then resampled using weights computed from those likelihoods.

If *X*_g,j_(*t*) denotes the collection of patient-level latent states within block *g* at time *t* for particle *j*, *Y*_g_(*t*) the corresponding observations, and *θ*_j_ the associated parameter vector for particle *j*, then the blockwise particle weight takes the form

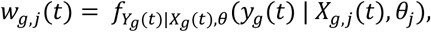

so that blocks contribute separately to the filtering update. This blockwise factorization is what allows the iterated filter to operate in a much higher-dimensional patient-level state space, where the latent dimension is driven by the number of patients rather than by a small set of aggregated units (36–38, 58). This iterative procedure drives the parameter ensemble toward regions of high likelihood while maintaining stochastic exploration early in the inference process. In BAIF, block membership is used to organize parameter sharing and likelihood evaluation. The latent transmission process remains patient-level, while inference is organized around block-specific transmission and importation parameters.

## Data, Materials, and Software Availability

Public sharing of the hospitalization records and microbiological surveillance data is not permitted under the data-use agreement with the participating hospital system. Code implementing BAIF an d demonstrating its use with synthetic data has been deposited at GitHub, https://github.com/jifanli/BAIF_synthetic_example.

## Acknowledgments

This research was supported by funding from National Institute of Allergy and Infectious Diseases R21AI180492 (N.N. and S.P.) and National Institute of General Medical Sciences R35GM156799 (S.P.). Portions of this work were conducted using the advanced computing resources provided by Texas A&M High Performance Research Computing (HPRC).

## Author Contributions

N.N. and S.P. designed research; J.L. performed research; J.L., Q.Y., S.P., and N.N. investigated results; J.L., Q.Y., S.P., and N.N. wrote the paper.

