## Supporting Information for "Inferring heterogeneous transmission and community introduction of antibiotic-resistant bacteria in hospital settings"

##### This PDF file includes:

Supporting text
Figs. S1 to S7
Tables S1 to S8
SI References

#### Supporting Information Text

### Hospitalization and Surveillance Data

Hospitalization records were converted to weekly patient-location histories for the period 2012–2016. For each week, patients present in the hospital system were indexed and linked to the ward in which they were located. These weekly patient sets define the time-varying co-location network used by the transmission model. Patients enter and leave the active network as they are admitted, discharged, and transferred, and the set of co-located neighbors is reconstructed from ward occupancy in each week. Within each weekly network, the colonization process is propagated through seven daily substeps.

A patient's importation event was placed at the first evidence of hospitalization during 2012–2016, defined as the earlier of the first ward-stay record and the first surveillance event. This rule produced one first-entry event for each of 354,100 patients; for 661 patients, the first surveillance event preceded the first ward-stay record. For auditing readmissions, hospitalization-account intervals for the same patient were also grouped into continuous observed hospital spells when their date ranges overlapped or were separated by no more than one calendar day. This separate audit identified 530,926 hospital spells, but later spells did not create additional importation events.

Microbiology records were converted into two matrices for each organism. The first matrix records observed positive detections, and the second records the testing schedule. These matrices have rows corresponding to patients and columns corresponding to the 261 weeks in 2012–2016. When an organism-specific culture date was recorded, a resistant culture was classified as positive and a susceptible culture as negative; a patient-week containing both was classified as positive. To retain organism-specific resistance results for which the corresponding culture date was missing, the observation week was assigned using another recorded culture date from the same hospitalization, or the hospitalization midpoint when no culture date was available. A surveillance patient-week without a matching ward-location record was retained as observed hospital presence but assigned no co-location neighbors. The observation model uses this testing schedule directly and does not assume that untested patient-weeks are negative.

The main text presents ward-level MRSA surveillance data. Figure S1 presents the corresponding ward-by-week heatmaps for LRPA, CRKP, and VRE. Figure S2 summarizes testing coverage and positive detections across all four organisms at the block level. These observed detection rates do not distinguish colonization present on admission from colonization acquired within the hospital.

### Ward Transfer Network and Block Construction

The ward-transfer network was reconstructed from hospitalization records. Nodes represent wards, and directed transfers were counted when a patient moved from one ward to another. For an ordered ward pair $(u, v)$, the directed transfer weight was

$$w_{uv} = \frac{\#\{\text{transfers from ward } u \text{ to ward } v\}}{\#\{\text{patients ever admitted to ward } u\}}.$$

Normalizing by the number of patients admitted to the source ward makes directed transfer intensity comparable across wards with different patient volumes. In the undirected network supplied to Infomap community detection (1), the edge between wards $u$ and $v$ was assigned weight $w_{uv} + w_{vu}$. The off-diagonal values of the resulting symmetric matrix were linearly rescaled to $[0, 1]$, yielding $W_{uv}$. Infomap was run using the `cluster_infomap` function in the R package `igraph`, with edge weights $W_{uv}^{0.08}$ and 10 trials.

The initial Infomap partition contained 10 communities. Each of the five communities without surveillance observations was merged into the retained community with which it had the greatest total edge weight, producing five movement-based ward blocks. The resulting partition was used as the spatial scale for estimating block-specific transmission rates and importation probabilities. The purpose of using blocks is not to remove patient movement from the model. Patient movement remains represented through the weekly co-location network. The blocks provide a feasible parameterization: wards in the same movement community share a transmission parameter and an importation probability, while patient movement determines the changing set of contacts through which transmission can occur.

Table S2 summarizes the five blocks. The ward counts are the numbers of wards assigned to each Infomap-derived block. Patient-ward stays count a patient once for each ward occupied during 2012–2016, so a patient who occupied multiple wards can contribute to multiple ward totals. The assigned-patient column reports the number of patients assigned to each fixed parameter block, with each patient assigned to the block containing the greatest number of their distinct recorded patient-days. Across the transfer records used to build the network, 84.2% of ward-to-ward transfers occurred between wards assigned to the same block. Figure S3 shows the complete block-to-block transfer matrix and compares the distribution of wards with the distribution of patient-ward stays across blocks.

For parameter assignment, recorded patient dates were summed directly within ward blocks, and each patient was assigned to the block containing the greatest number of distinct recorded patient-days. Multiple recorded locations in the same block on the same date contributed one block-day. Ties in block-days were resolved by the number of patient-date-location records in the tied blocks and then, if needed, by the lower block number.

#### Agent-Based Colonization Model

**Purpose.** The model represents colonization acquisition, clearance, and importation for individual patients over the study period. Recorded hospitalization histories determine hospital presence and co-location, while the surveillance schedule determines which patient-weeks contribute observations. BAIF uses both the latent colonization process and the observation model to estimate heterogeneous transmission and importation parameters.

**Entities, state variables, and scales.** Patients are the model entities, with $X_i(t) \in \{\mathcal{S}, \mathcal{C}\}$ denoting patient $i$'s binary latent colonization state, where $\mathcal{S}$ and $\mathcal{C}$ denote susceptible and colonized, respectively. Hospital presence and ward location are time-varying observed attributes reconstructed from hospitalization records. The latent colonization process is represented at the patient level, contacts are defined by contemporaneous ward co-location, and transmission and importation parameters vary across five ward blocks. Co-location networks are updated and surveillance observations are evaluated weekly, with colonization propagated through seven daily substeps within each week.

**Process overview and scheduling.** At the start of a week, the active patient set and ward co-location network are updated from hospitalization and surveillance records. On each patient's first observed day in the study, colonization probability is initialized from the block-specific importation probability. Patients present without a co-located neighbor remain in the active set; their neighbor set is empty, so they undergo clearance but cannot acquire colonization through co-location during that week. Transmission and clearance are propagated for seven daily substeps. At the end of the observation week, simulated detections are compared with the recorded tests using blockwise likelihoods, followed by blockwise resampling during inference.

**Design concepts.** Interaction is represented by contemporaneous ward co-location, which captures opportunities for direct contact and indirect transmission mediated by shared healthcare workers, equipment, or ward environments without assigning those routes separate parameters (2–5). Heterogeneity enters through block-specific transmission and importation parameters. Partial observation is represented by the recorded testing schedule and imperfect diagnostic sensitivity.

**Initialization.** The importation probability is applied once, on a patient's first observed study day. Thereafter, the state is retained across discharge and readmission; clearance continues during observed and unobserved hospital-absence periods, and readmission does not trigger another importation reset. Detailed initialization and continuation rules are given below.

**Input data.** Exogenous inputs are weekly patient presence reconstructed from hospitalization and surveillance records, ward location reconstructed from hospitalization records, the resulting co-location networks, the organism-specific testing and detection matrices, fixed clearance and diagnostic-sensitivity values, and ward-block membership.

**Patient states and importation.** Let $X_i(t) \in \{\mathcal{S}, \mathcal{C}\}$ denote the latent state of patient $i$ at daily substep $t$. The simulator propagates $S_i(t) = P\{X_i(t) = \mathcal{S}\}$ and $C_i(t) = P\{X_i(t) = \mathcal{C}\}$, with $S_i(t) + C_i(t) = 1$. Let $g(i)$ denote the fixed block assigned to patient $i$ for parameterization. At patient $i$'s first observed day $t_e$ in the study, the colonization probability is initialized as

$$C_i(t_e) = C_{0,g(i)}.$$

**Transmission and clearance.** After admission, susceptible patients can acquire colonization through contemporaneous ward co-location with colonized patients. Let $N_i(t)$ be the set of co-located neighbors of patient $i$. The acquisition hazard is

$$\lambda_i(t) = \beta_{g(i)} \sum_{j \in N_i(t)} C_j(t),$$

where $\beta_{g(i)}$ is the transmission rate for the assigned block. The daily update is

$$C_i(t+1) - C_i(t) = \lambda_i(t) S_i(t) - \alpha C_i(t).$$

This expression is the discrete-time update implemented by the simulator. For each weekly co-location network, the compiled transmission routine applies this update seven times, once for each daily substep, before the network is replaced by the following week's network. The clearance parameter $\alpha$ is held fixed by organism (Table S1), while $\beta_g$ and $C_{0,g}$ are estimated.

**Observation model.** The observation model uses the recorded testing schedule. Let $T_i(w)$ indicate whether patient $i$ was tested during week $w$, and let $Y_i(w)$ denote the observed detection. Untested patient-weeks are excluded from the observation likelihood. For tested patient-weeks, the model assumes diagnostic sensitivity $p_{\text{sens}} = 0.4$ and specificity 1:

$$P\{Y_i(w) = 1 \mid X_i(w) = \mathcal{C}, T_i(w) = 1\} = p_{\text{sens}}, \qquad P\{Y_i(w) = 1 \mid X_i(w) = \mathcal{S}, T_i(w) = 1\} = 0.$$

In the filtering likelihood, observations are compared at the block-week level. If $A_g(w)$ is the set of tested patients in block $g$ at week $w$, the expected number of detections for particle $j$ is

$$\mu_{g,j}(w) = p_{\text{sens}} \sum_{i \in A_g(w)} C_{i,j}(w).$$

The observed block-week count $y_g(w)$ is compared with $\mu_{g,j}(w)$ using a scaled-Poisson working objective. With $z_g(w) = p_o y_g(w)$, $m_{g,j}(w) = p_o \mu_{g,j}(w)$, and $p_o = 10$, its log value is

$$\ell_{g,j}(w) = z_g(w) \log m_{g,j}(w) - m_{g,j}(w) - \log \Gamma\{z_g(w) + 1\},$$

with the usual zero-count limit. For the integer counts in the hospital data, this is the Poisson log mass after multiplying both the observed and expected counts by $p_o$. For deterministic noninteger targets in the synthetic validation, the gamma-function expression evaluates the same working objective without rounding. Conditional on the target count, the objective is maximized when the expected count equals that target. The multiplier $p_o = 10$ makes log-objective contrasts ten times those obtained with $p_o = 1$ and is used throughout the analysis.

#### Blockwise Agent-Based Iterated Filtering

**Particle representation and notation.** BAIF represents uncertainty with an ensemble of state–parameter particles. Each particle carries susceptible and colonized probabilities for all patients together with block-specific transmission and importation parameters. Superscript $P$ denotes the pre-filter stage, in which parameters have been perturbed and patient states have been advanced through the current week's ABM transition but have not yet been weighted or resampled; superscript $F$ denotes the state and parameters after blockwise likelihood weighting and resampling. The index $m = 1, \ldots, M$ denotes filtering iterations, $n = 1, \ldots, N$ denotes observation weeks, and $j = 1, \ldots, J$ denotes particles. At week $n$, let $\mathcal{K}_n = \{\kappa_{n,1}, \ldots, \kappa_{n,G_n}\}$ denote the partition used for blockwise filtering on the current network, and let $y_n^{\kappa_{n,g}}$ denote the observations associated with filtering block $\kappa_{n,g}$. This notation allows the filtering partition to vary with the agent set and contact structure. In the present analysis, patient membership in the five parameter blocks is fixed within each annual fit, so $G_n = 5$ and $\kappa_{n,g} = K_g$ for every week, whereas the active patient set and contact network vary with $n$.

Let $X_{n,j}^{F,m}$ contain the susceptible and colonized probabilities for every patient after filtering week $n$. The corresponding logit-scale parameter component for filtering block $\kappa_{n,g}$ is

$$\theta_{n,j}^{\kappa_{n,g},F,m} = \left\{\operatorname{logit}(\beta_{g,n,j}^{F,m}), \operatorname{logit}(C_{0,g,n,j}^{F,m})\right\}.$$

**Perturbation and cooling.** At the start of iteration $m$, the parameter swarm is inherited from the end of iteration $m-1$, whereas the latent states are reset to the fixed initial particle states for that calendar year. At each observation week, Gaussian random-walk perturbations are applied to the block parameters on the logit scale before patient-state propagation (6–8). With the geometric cooling used in the final analyses, the perturbation multiplier is

$$a_{m,n} = 0.5^{\{m-1+n/N\}/50}.$$

Before multiplication by $a_{m,n}$, the block-level random-walk standard deviations are 0.045 for $\operatorname{logit}(\beta_g)$ and 0.025 for $\operatorname{logit}(C_{0,g})$. Perturbations are independent across blocks and particles, and all patients assigned to a block share the resulting $\beta_g$ and $C_{0,g}$ values.

**Full-network propagation and blockwise filtering.** Let $\mathcal{A}_n$ denote the active patient set in week $n$. For every particle, patients in $\mathcal{A}_n$ may acquire colonization through the weekly co-location network $\mathcal{H}_n$ and may clear colonization, whereas patients who were present in an earlier week but are outside $\mathcal{A}_n$ undergo clearance without co-location exposure. Transmission can occur between co-located patients assigned to different parameter blocks. After this state update, likelihoods are evaluated and resampling is performed separately within each block. If $\mathcal{T}_{n,g} = \{i \in \kappa_{n,g} : T_i(n) = 1\}$ is the set of tested patients in filtering block $\kappa_{n,g}$, then

$$y_g(n) = \sum_{i \in \mathcal{T}_{n,g}} Y_i(n), \qquad \mu_{n,j}^{\kappa_{n,g},m} = p_{\text{sens}} \sum_{i \in \mathcal{T}_{n,g}} C_{i,n,j}^{P,m},$$

and the corresponding scaled-Poisson log objective is

$$\ell_{n,j}^{\kappa_{n,g},m} = p_o y_g(n) \log\left\{p_o \mu_{n,j}^{\kappa_{n,g},m}\right\} - p_o \mu_{n,j}^{\kappa_{n,g},m} - \log\Gamma\{p_o y_g(n) + 1\},$$

with the usual zero-count limit. The numerically stabilized block weight is

$$w_{n,j}^{\kappa_{n,g},m} = \exp\left\{\ell_{n,j}^{\kappa_{n,g},m} - \max_{1 \le r \le J} \ell_{n,r}^{\kappa_{n,g},m}\right\}, \qquad p_{\text{sens}} = 0.4, \quad p_o = 10.$$

When $\mathcal{T}_{n,g}$ is empty, $w_{n,j}^{\kappa_{n,g},m} = 1$ for every particle. The subtracted offset is restored when the block-filter likelihood is calculated. Using these weights, each block independently resamples its state and parameter components from the particle ensemble. The resampled components from all blocks are then recombined to form the new full-system particles. A standard global particle filter would use all observations to update each full hospital-wide particle. In a high-dimensional state space, small differences in likelihood contributions accumulate in a global particle weight, which can concentrate the normalized weights on very few particles and make an adequate particle representation computationally prohibitive (6). BAIF instead bases each block correction on that block's observations and recombines the corrected components, so that no single particle weight accumulates likelihood differences across all blocks, while cross-block transmission is preserved during state propagation.

**Initialization.** The importation probability $C_{0,g}$ initializes each patient's state on the first observed day during 2012–2016 and is not reapplied at readmission. For 2013–2016, each chain continues from its own terminal patient-state ensemble from the preceding year, rather than that of another chain with a higher likelihood.

For each organism, the real-data analysis used 100 inference chains initialized from the organism-specific grid in Table S1. Initial $\beta_g$ values are logarithmically spaced over the reported range, initial $C_{0,g}$ values are linearly spaced, and each chain begins with the same initial value in all five blocks. The two 100-point grids are paired by a deterministic permutation constrained to have an absolute Spearman rank correlation no greater than 0.05.

**Initial-range calibration.** Before the 100-chain analysis, the organism-specific starting scales were examined using a deterministic diagnostic based on the 2012 data. For each candidate pair $(\beta, C_0)$, all five blocks were temporarily assigned the same values, the agent-based model was propagated on the observed 2012 co-location networks and testing schedule, and the expected block-week detections were compared with the observed counts using the scaled-Poisson objective above. The candidate $\beta$ ranges were $10^{-6}$–0.008 for MRSA, $10^{-4}$–0.025 for LRPA, $10^{-6}$–0.004 for CRKP, and $10^{-12}$–0.03 for VRE; the corresponding $C_0$ ranges were 0.10–0.22, 0.04–0.16, 0.03–0.10, and 0.10–0.24. The objective-maximizing grid pairs $(\beta, C_0)$ were $(0.002, 0.14)$ for MRSA, $(0.006, 0.10)$ for LRPA, $(0.0005, 0.05)$ for CRKP, and $(10^{-12}, 0.203)$ for VRE. At $C_0 = 0.203$, the VRE objective differed by 0.075 log-objective units between $\beta = 10^{-12}$ and $10^{-5}$, indicating a near-zero ridge rather than a precisely resolved optimum. This common-parameter diagnostic was used only to determine organism-specific starting scales; it was not a block-specific fit or a BAIF iteration. The initial ranges in Table S1 were deliberately placed above the diagnostic maxima so that convergence could be evaluated from elevated starting values, while the BAIF analysis allowed the five block-specific parameters to evolve independently.

**Pseudocode.**

**Algorithm S1** The BAIF algorithm

**Input:** week-indexed ABM transition simulators $\mathcal{F}_{1:N}$, blockwise observation evaluators $\mathcal{G}_{1:N}$, and data $y_{1:N}$;
active-agent sets $\mathcal{A}_{1:N}$, dynamic contact networks $\mathcal{H}_{1:N}$, and filtering partitions $\mathcal{K}_{1:N}$;
number of iterations $M$, number of particles $J$, and parameter perturbation kernels $\mathcal{Q}_{m,n}$;
initial joint state–parameter ensemble $\{(X_{0,j}, \theta^0_j)\}_{j=1}^J$.

**For** $m$ in $1:M$
  Set $\theta^{F,m}_{0,j} \leftarrow \theta^{m-1}_j$ and $X^{F,m}_{0,j} \leftarrow X_{0,j}$ for $j = 1, \dots, J$.
  **For** $n$ in $1:N$
    Update the active-agent set to $\mathcal{A}_n$, the contact network to $\mathcal{H}_n$, and the filtering partition to $\mathcal{K}_n = \{\kappa_{n,g}\}_{g=1}^{G_n}$.
    **For** $\kappa_{n,g} \in \mathcal{K}_n$
      Draw $\theta^{\kappa_{n,g},P,m}_{n,j} \sim \mathcal{Q}^{\kappa_{n,g}}_{m,n}(\cdot \mid \theta^{F,m}_{n-1,j})$ for $j = 1, \dots, J$.
    **End For**
    Recombine $\theta^{P,m}_{n,j} \leftarrow (\theta^{\kappa_{n,g},P,m}_{n,j})_{\kappa_{n,g} \in \mathcal{K}_n}$ for every particle $j$.
    **For** $j$ in $1:J$
      Propagate the full agent-level state with the ABM transition simulator:
      $X^{P,m}_{n,j} \leftarrow \mathcal{F}_n\{X^{F,m}_{n-1,j}; \mathcal{A}_n, \mathcal{H}_n, \theta^{P,m}_{n,j}\}$.
    **End For**
    **For** $\kappa_{n,g} \in \mathcal{K}_n$
      **For** $j$ in $1:J$
        Compute $w^{\kappa_{n,g},m}_{n,j} \leftarrow \mathcal{G}_{n,g}\{y^{\kappa_{n,g}}_n; X^{\kappa_{n,g},P,m}_{n,j}, \theta^{\kappa_{n,g},P,m}_{n,j}\}$.
      **End For**
      Draw $s^{\kappa_{n,g},m}_{n,j}$ independently for $j = 1, \dots, J$, with $\Pr(s^{\kappa_{n,g},m}_{n,j} = r) = w^{\kappa_{n,g},m}_{n,r} / \sum_{q=1}^J w^{\kappa_{n,g},m}_{n,q}$.
      Set $X^{\kappa_{n,g},F,m}_{n,j} \leftarrow X^{\kappa_{n,g},P,m}_{n,s^{\kappa_{n,g},m}_{n,j}}$ and $\theta^{\kappa_{n,g},F,m}_{n,j} \leftarrow \theta^{\kappa_{n,g},P,m}_{n,s^{\kappa_{n,g},m}_{n,j}}$ for $j = 1, \dots, J$.
    **End For**
    Recombine the filtered blocks to form each new full-system particle:
    $X^{F,m}_{n,j} \leftarrow (X^{\kappa_{n,g},F,m}_{n,j})_{\kappa_{n,g} \in \mathcal{K}_n}$ for $j = 1, \dots, J$.
    $\theta^{F,m}_{n,j} \leftarrow (\theta^{\kappa_{n,g},F,m}_{n,j})_{\kappa_{n,g} \in \mathcal{K}_n}$ for $j = 1, \dots, J$.
    Set $\widehat{\ell}^m_{b,n} \leftarrow \sum_{\kappa_{n,g} \in \mathcal{K}_n} \log\{J^{-1} \sum_{j=1}^J w^{\kappa_{n,g},m}_{n,j}\}$.
  **End For**
  Set $\theta^m_j \leftarrow \theta^{F,m}_{N,j}$ for $j = 1, \dots, J$ and $\widehat{\ell}^m_b \leftarrow \sum_{n=1}^N \widehat{\ell}^m_{b,n}$.
**End For**

**Output:** Final joint state–parameter ensemble $\{(X^{F,M}_{N,j}, \theta^M_j)\}_{j=1}^J$ and final-iteration block-filter likelihood $\widehat{\ell}^M_b$.

**Implementation settings.** The BAIF workflow used for the real-data analysis was:

1. For each organism, create five batches of 20 inference chains, giving 100 chains.
2. Within each chain and year, initialize $\beta_g$ and $C_{0,g}$ from the organism-specific starting grid in Table S1.
3. Run BAIF for 15 filtering iterations with 30 particles.

4. At the end of a year, save the terminal susceptible and colonized state particles for each chain.

5. Initialize the same chain in the next year from its saved terminal state particles, rather than using the single highest-likelihood chain for all following years.

6. After 2016, combine the yearly replicate estimates and compute each chain's five-year mean parameter value for each organism-block combination.

Random-number streams were set deterministically by organism, batch, and year. Parallel chain-level computations used `doRNG` so that repeated runs with the same inputs reproduce the same draws. The synthetic-validation streams and initial-grid pairing were likewise set deterministically.

#### Synthetic Validation Design

The synthetic validation asks whether BAIF can recover known block-specific transmission and importation parameters from partially observed data generated under the same agent-based model and recorded testing schedule. Synthetic observations were generated using the 2012 patient set and ward co-location networks together with the organism-specific testing schedule and fixed $\alpha$ value associated with each parameter setting. The validation used 20 independent inference realizations, 30 particles, and 15 BAIF iterations. Iteration 0 in the convergence figures is the initial parameter distribution before BAIF updating.

Panel A of main-text Fig. 3 shows convergence for settings A–C after scaling every trajectory by its block-specific true value, and panel B compares the final estimates with the generating values across settings A–D. Figure S4 gives a detailed block-specific convergence example for setting D. Within each setting, the block-specific transmission and importation values were paired at random, allowing high values of the two parameters to occur in either the same or different blocks. Initial values were deliberately set above the true values so that convergence is visible as movement toward the horizontal reference lines. For each setting, the 20 initial $\beta_g$ values are logarithmically spaced from 1.35 to 6.75 times the largest true $\beta_g$. The initial $C_{0,g}$ values are linearly spaced from 0.04 above the largest true $C_{0,g}$ to a setting-specific upper bound no greater than 0.95. Table S5 gives the generating values together with the medians and 2.5th–97.5th percentile intervals of the final estimates across the 20 synthetic inference realizations.

Among the 20 setting-block values for each parameter in main-text Fig. 3B, the 2.5th–97.5th percentile intervals contained the corresponding generating value for 16 $\beta_g$ values and 19 $C_{0,g}$ values. Figure S5 shows the two parameters jointly: individual estimates from each inference realization are plotted relative to their generating values, and the block-specific medians remain concentrated near the generating parameter pairs. Together, main-text Fig. 3 and Fig. S5 show that BAIF recovers distinct block-specific transmission and importation values across the four synthetic settings.

For each block and week in the synthetic validation, the observation was set to the sum of the model-implied detection probabilities among tested patients, $p_{\text{sens}} \sum_i C_i(w)$. Thus, the validation used a deterministic, possibly noninteger observation target rather than sampling additional patient-level Bernoulli detection outcomes. This choice removes extra observation noise so that the validation tests whether the algorithm can distinguish $\beta_g$ from $C_{0,g}$, rather than whether a single noisy realization happens to favor parameter values away from the generating truth.

#### Summaries of Real-Data Parameter Estimates

For Fig. 4A in the main text, the five yearly estimates from 2012–2016 were averaged for each inference chain and pathogen-block pair. The boxplots then summarize the distribution of these five-year chain averages across 100 chains. Table S6 gives the corresponding medians, interquartile ranges, and central 95% intervals for both $\beta_g$ and $C_{0,g}$. Figure S6 reports the percentage of chains in which each block has the largest five-year mean parameter estimate for an organism. Figure S7 compares the all-chain medians with medians from the 20 chains having the highest mean likelihood.

#### Fitted-Model Simulations and Contribution Decomposition

For Fig. 4B, one colonization-probability trajectory was propagated from the fitted yearly parameters in each of the 100 inference chains. The simulator records the expected number of colonized tested patients each week. To represent observed detections under imperfect diagnostic sensitivity, each chain-week expectation was multiplied by $p_{\text{sens}} = 0.4$ and converted to a count distribution by Poisson sampling. For the plotted simulation intervals, 500 independent Poisson draws were generated from each of the 100 chain-week expectations, yielding 50,000 observation draws per organism-week. The orange line is the median across these draws, and the shaded regions are their central 50% and 95% intervals. Coverage was computed as the proportion of observed weekly detections falling within the central 95% simulation interval.

For Fig. 5, the full fitted-model simulation was compared with an importation-only simulation. The importation-only simulation used the same fitted $C_{0,g}$ values but set $\beta_g = 0$. For each organism and block, expected colonized patient-weeks were accumulated over patients active in each week. The importation contribution is the mean of the 100 importation-only totals, one obtained from each fitted inference chain, and the transmission contribution is the difference between the means of the corresponding full-model and importation-only totals. Each quantity is divided by the block's total active patient-weeks over 2012–2016 and multiplied by 1,000, yielding the contributions per 1,000 active patient-weeks shown in the main text.

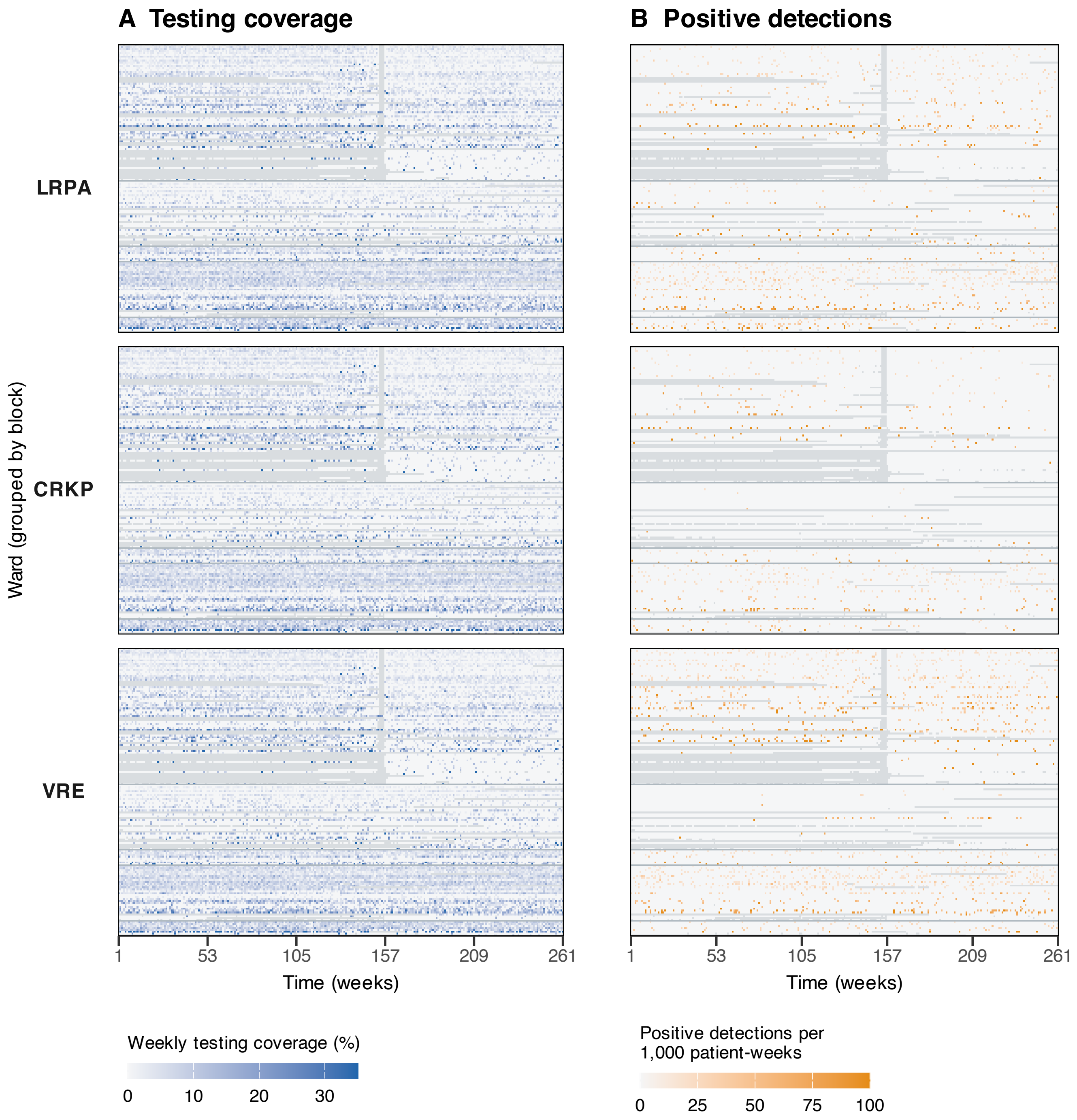


**Fig. S1.** Ward-level surveillance for LRPA, CRKP, and VRE, 2012–2016. (A) Weekly testing coverage, calculated as the percentage of patient-weeks with a test for the corresponding organism among all patient-weeks present in each ward. (B) Positive detections per 1,000 patient-weeks. Wards with at least 500 patient-weeks are grouped by block and ordered within each block by decreasing total patient-weeks over 2012–2016. Gray cells indicate ward-weeks with no patients present. For visualization, the color scales are capped at 35% testing coverage and 100 detections per 1,000 patient-weeks; values exceeding these thresholds are assigned the maximum scale color.

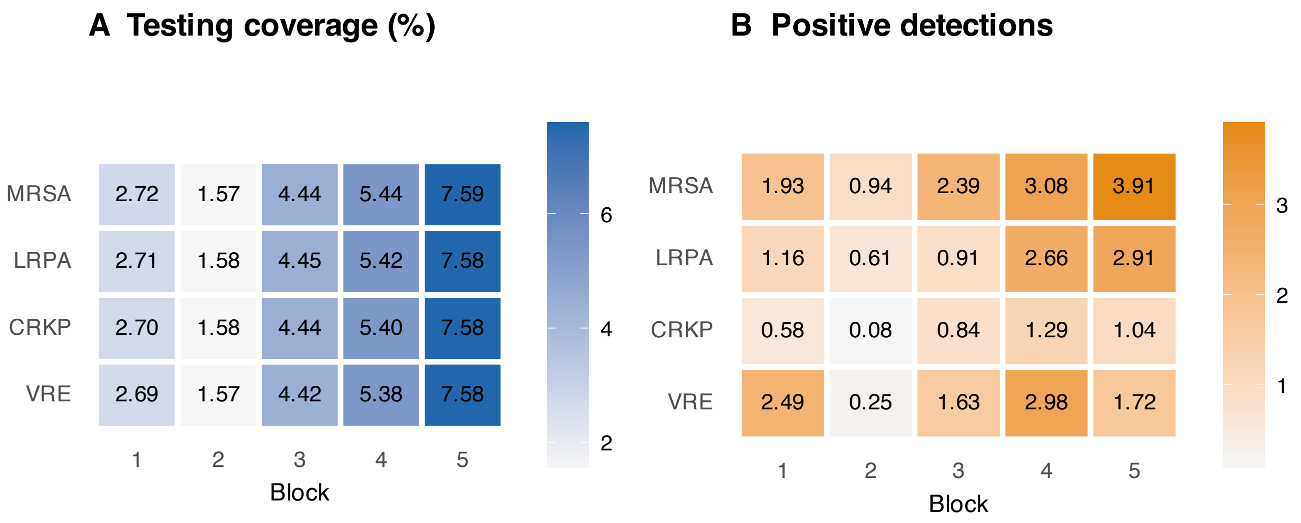


**Fig. S2.** Block-level surveillance rates for all four organisms, 2012–2016. (A) Percentage of patient-weeks with a test for the corresponding organism among all patient-weeks present in each block. (B) Positive detections per 1,000 patient-weeks.

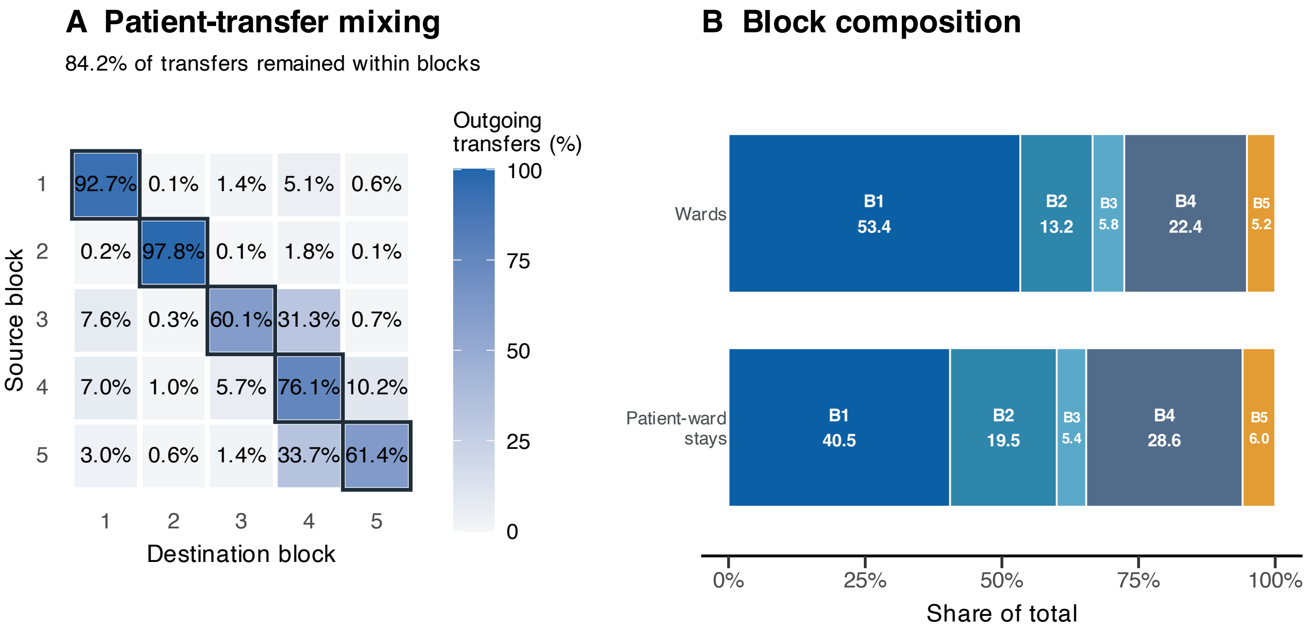


**Fig. S3.** Diagnostics for the five-block ward partition. (A) Row-normalized patient-transfer matrix. For each patient, ward records were ordered by date. A directed transition from ward $u$ to ward $v$ was recorded when successive records listed different wards; repeated occurrences of the same ordered ward pair for that patient were counted once. Cells give the percentage of transitions from each source block entering each destination block. Outlined diagonal cells are within-block transitions. Summed over source blocks, 84.2% of transitions occurred between wards assigned to the same block. (B) Distribution of wards and patient-ward stays across blocks. Patient-ward stays count a patient once in each ward occupied during 2012–2016. Labels give block number and percentage of the corresponding total.

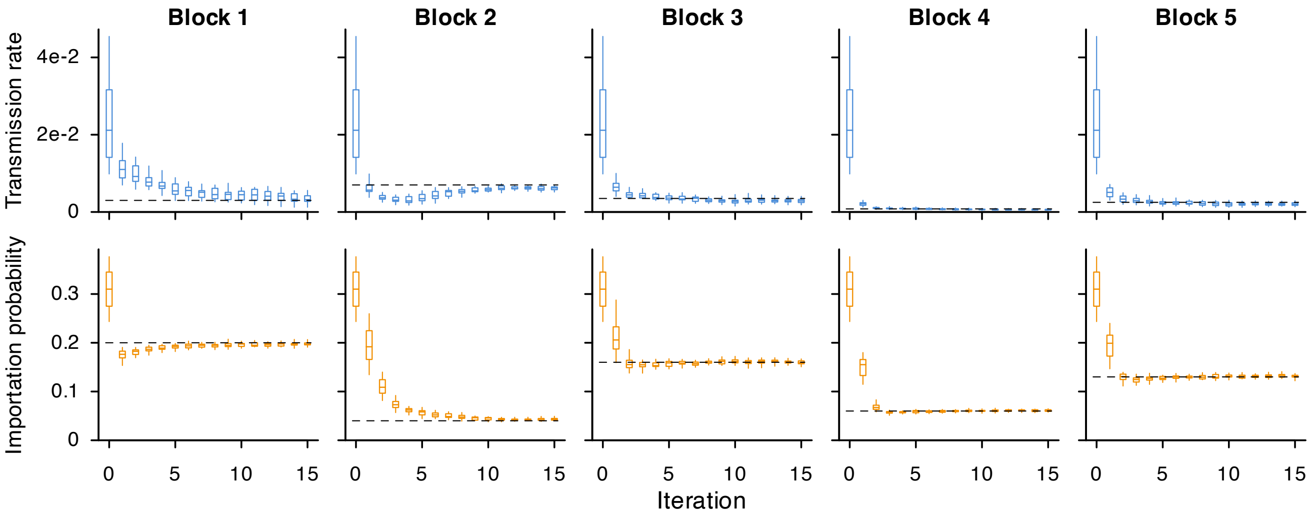


**Fig. S4.** Recovery of block-specific parameters from synthetic setting D using BAIF. Columns correspond to the five ward blocks, and iteration 0 shows the initial parameter distributions before BAIF updating. The top row presents block-specific transmission rates, $\beta_g$, and the bottom row presents block-specific importation probabilities, $C_{0,g}$. Boxes indicate the interquartile range and whiskers the central 95% interval across 20 independent BAIF inference realizations. Horizontal dashed lines mark the true parameter values used to generate the synthetic outbreak. Convergence of the particle distributions toward these reference values demonstrates recovery of both transmission and importation parameters under partial observation.

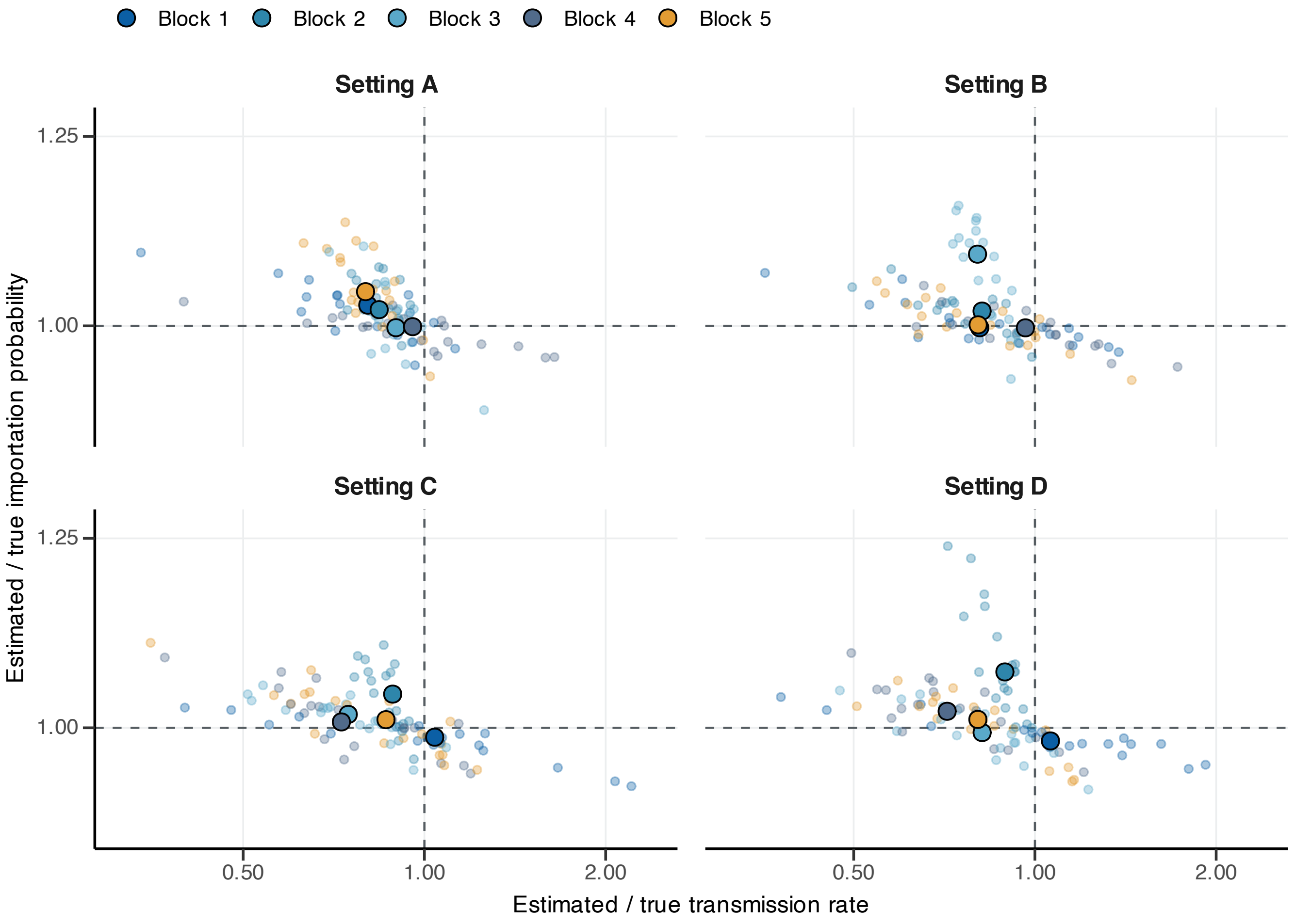


**Fig. S5.** Paired final estimates of transmission and importation parameters across synthetic settings A–D. Each small point is the final estimate from one of 20 independent BAIF inference realizations for one block, divided by the corresponding generating value. Large outlined points are block-specific medians across the 20 realizations. Dashed lines intersect at the generating parameter pair $(1, 1)$. The concentration of the block-specific median pairs near $(1, 1)$ shows simultaneous recovery of the transmission and importation values. Both axes are logarithmic.

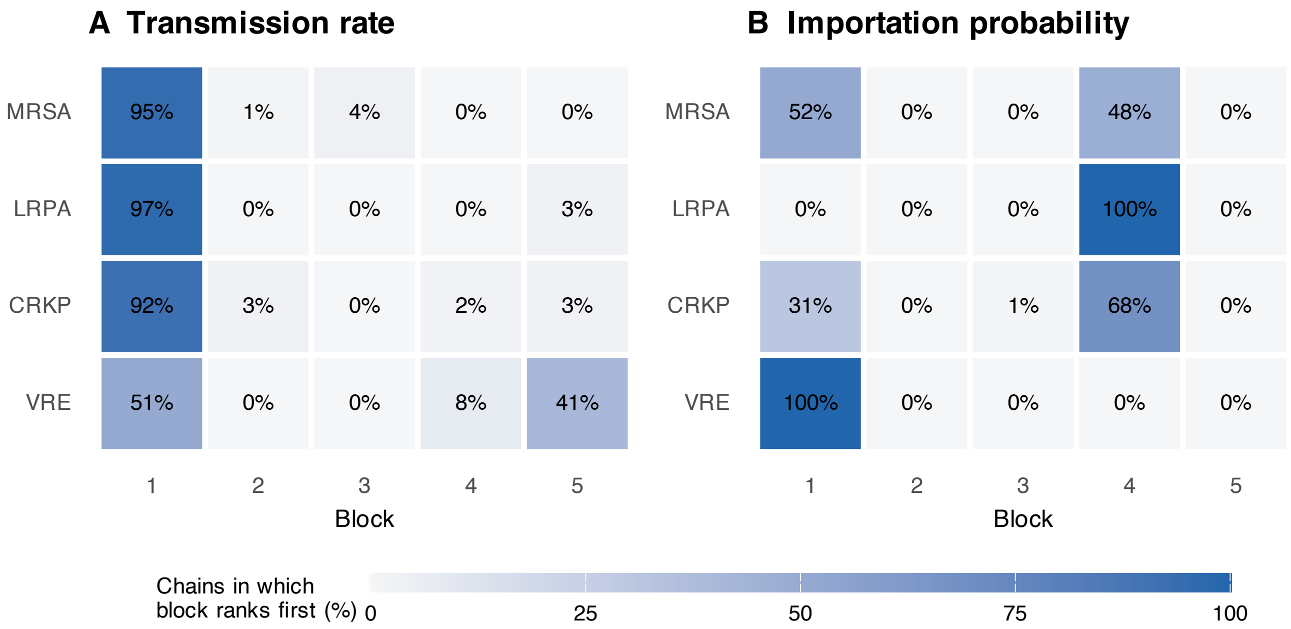


**Fig. S6.** Frequency of each block ranking highest across 100 BAIF inference chains. (A) Transmission rates. (B) Importation probabilities. Each cell is the percentage of chains in which the indicated block has the largest five-year mean estimate for that organism.

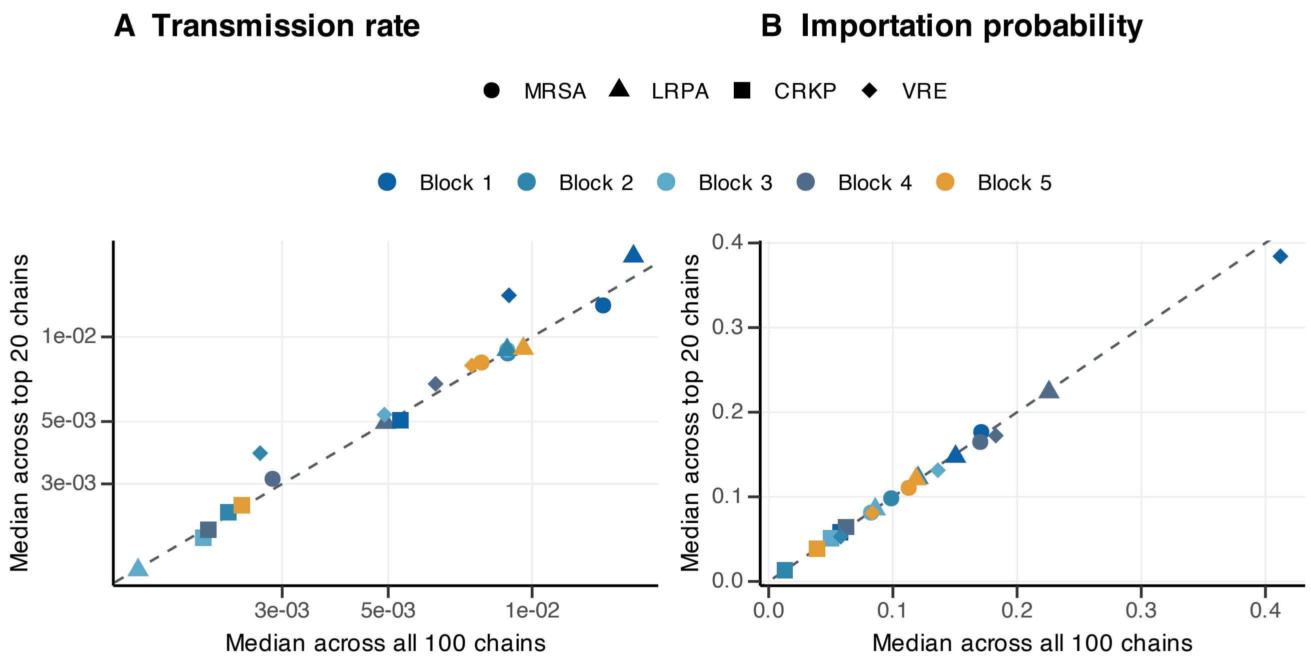


**Fig. S7.** Comparison of block-specific parameter estimates using all 100 inference chains and the 20 chains with the highest mean likelihood. (A) Transmission rates. (B) Importation probabilities. Horizontal coordinates are medians of the five-year chain-average estimates across all 100 chains; vertical coordinates are medians after retaining the 20 chains with the highest mean likelihood for each organism. Dashed lines are identity lines, colors identify blocks, and shapes identify organisms.

**Table S1. Organism-specific surveillance totals, clearance parameters, and inference initialization ranges. Tested patient-weeks and positive detections are totals from the binary weekly surveillance matrices for 2012–2016. With daily clearance probability $\alpha$, the reported median duration is $\log(0.5)/\log(1-\alpha)$, obtained by setting the probability of remaining colonized after $d$ days, $(1-\alpha)^d$, equal to 0.5. The fixed clearance values were chosen to fall within published duration ranges for the corresponding organisms or related resistant phenotypes (9–12). The initial ranges are the organism-specific starting grids used for real-data BAIF refits.**

| Organism | Tested patient-weeks | Positive detections | $\alpha$ | Median duration (days) | Initial $\beta_g$ range | Initial $C_{0,g}$ range |
|---|---|---|---|---|---|---|
| MRSA | 34,759 | 2,110 | 0.003 | 230.7 | 0.004–0.016 | 0.18–0.30 |
| LRPA | 34,672 | 1,480 | 0.016 | 43.0 | 0.010–0.040 | 0.14–0.26 |
| CRKP | 34,549 | 695 | 0.003 | 230.7 | 0.001–0.006 | 0.08–0.16 |
| VRE | 34,426 | 2,038 | 0.017 | 40.4 | 0.0005–0.005 | 0.24–0.36 |

**Table S2. Numbers of wards, patient-ward stays, and assigned patients in each block. Patient-ward stays are the block totals displayed in the block-level network panel. Assigned patients are the patient counts in the block assignment file used by the continuous BAIF analyses.**

| Block | Wards | Patient-ward stays | Share of patient-ward stays | Assigned patients |
|---|---|---|---|---|
| 1 | 1,032 | 422,834 | 40.5% | 165,568 |
| 2 | 256 | 203,549 | 19.5% | 81,910 |
| 3 | 112 | 56,313 | 5.4% | 19,079 |
| 4 | 434 | 298,526 | 28.6% | 70,627 |
| 5 | 100 | 62,213 | 6.0% | 16,916 |

**Table S3. Diagnostics for the fixed patient-block assignment. Stay-row counts refer to records considered before collapsing them to distinct patient dates for the primary cumulative block-day rule. Rows without either a mapped room or a mapped ward location were excluded from the duration calculation; every patient retained in the analysis had at least one mapped record and received a fixed block.**

| Quantity | Value |
|---|---|
| Patients assigned to a fixed block | 354,100 |
| Patients tied on the maximum block-day total | 5,513 |
| Stay rows assigned through room membership | 3,361,560 |
| Stay rows assigned through ward-location fallback | 480 |
| Stay rows without a mapped room or ward location | 9 |

**Table S4. Computational settings for the final BAIF analyses. The synthetic validation and real-data refits use the same BAIF model code, particle count, iteration count, random-walk perturbations, and scaled-Poisson objective.**

| Analysis | Data context | Chains or replicates | Particles | BAIF iterations | Main outputs |
|---|---|---|---|---|---|
| Synthetic validation | 2012 observed movement and testing schedule | 20 inference chains per setting | 30 | 15 | Parameter trajectories and joint true-value recovery, Fig. 3 and Figs. S4 and S5 |
| Real-data parameter estimation | 2012–2016 observed data, run separately by organism and year | 100 chains per organism | 30 | 15 per year | Yearly estimates, five-year chain means, Fig. 4A |
| Fitted-model simulations | 2012–2016 fitted yearly parameters | 100 fitted chains per organism | N/A | N/A | Weekly detection intervals, coverage, Fig. 4B |
| Contribution decomposition | Full and importation-only fitted-model simulations | 100 fitted chains per organism | N/A | N/A | Block importation and transmission contributions, Fig. 5 |

**Table S5. Generating values and synthetic-validation recovery summaries. Values are final medians with central 95% intervals across 20 independent BAIF inference realizations. Main-text Fig. 3A shows convergence for settings A–C, and Fig. 3B shows final calibration across settings A–D; Fig. S4 shows detailed convergence for setting D, and Fig. S5 summarizes joint recovery across all four settings.**

| Setting | Block | True $\beta_g$ | Final $\beta_g$ median (95% interval) | True $C_{0,g}$ | Final $C_{0,g}$ median (95% interval) | $\alpha$ |
|---|---|---|---|---|---|---|
| A | 1 | $7.00 \times 10^{-3}$ | $5.64 \times 10^{-3}$ ($3.14 \times 10^{-3}$–$7.58 \times 10^{-3}$) | 0.05 | 0.051 (0.048–0.054) | 0.016 |
| A | 2 | $6.00 \times 10^{-3}$ | $5.05 \times 10^{-3}$ ($4.50 \times 10^{-3}$–$5.62 \times 10^{-3}$) | 0.18 | 0.183 (0.178–0.193) | 0.016 |
| A | 3 | $3.50 \times 10^{-3}$ | $3.14 \times 10^{-3}$ ($2.59 \times 10^{-3}$–$3.95 \times 10^{-3}$) | 0.09 | 0.090 (0.084–0.099) | 0.016 |
| A | 4 | $5.00 \times 10^{-4}$ | $4.78 \times 10^{-4}$ ($2.56 \times 10^{-4}$–$8.09 \times 10^{-4}$) | 0.22 | 0.220 (0.212–0.226) | 0.016 |
| A | 5 | $4.00 \times 10^{-3}$ | $3.19 \times 10^{-3}$ ($2.63 \times 10^{-3}$–$4.04 \times 10^{-3}$) | 0.07 | 0.073 (0.067–0.078) | 0.016 |
| B | 1 | $4.00 \times 10^{-3}$ | $3.24 \times 10^{-3}$ ($1.76 \times 10^{-3}$–$5.41 \times 10^{-3}$) | 0.10 | 0.100 (0.097–0.106) | 0.003 |
| B | 2 | $2.50 \times 10^{-3}$ | $2.04 \times 10^{-3}$ ($1.34 \times 10^{-3}$–$2.41 \times 10^{-3}$) | 0.24 | 0.244 (0.233–0.254) | 0.003 |
| B | 3 | $3.00 \times 10^{-3}$ | $2.41 \times 10^{-3}$ ($2.21 \times 10^{-3}$–$2.74 \times 10^{-3}$) | 0.04 | 0.044 (0.038–0.046) | 0.003 |
| B | 4 | $4.00 \times 10^{-4}$ | $3.86 \times 10^{-4}$ ($2.58 \times 10^{-4}$–$6.17 \times 10^{-4}$) | 0.15 | 0.150 (0.143–0.156) | 0.003 |
| B | 5 | $2.00 \times 10^{-3}$ | $1.61 \times 10^{-3}$ ($1.11 \times 10^{-3}$–$2.61 \times 10^{-3}$) | 0.20 | 0.200 (0.190–0.210) | 0.003 |
| C | 1 | $4.00 \times 10^{-3}$ | $4.16 \times 10^{-3}$ ($1.75 \times 10^{-3}$–$8.57 \times 10^{-3}$) | 0.23 | 0.227 (0.215–0.235) | 0.017 |
| C | 2 | $8.00 \times 10^{-3}$ | $7.09 \times 10^{-3}$ ($6.16 \times 10^{-3}$–$8.10 \times 10^{-3}$) | 0.08 | 0.083 (0.078–0.088) | 0.017 |
| C | 3 | $4.50 \times 10^{-3}$ | $3.36 \times 10^{-3}$ ($2.31 \times 10^{-3}$–$4.62 \times 10^{-3}$) | 0.19 | 0.193 (0.183–0.199) | 0.017 |
| C | 4 | $8.00 \times 10^{-4}$ | $5.82 \times 10^{-4}$ ($3.73 \times 10^{-4}$–$9.43 \times 10^{-4}$) | 0.05 | 0.050 (0.048–0.054) | 0.017 |
| C | 5 | $3.00 \times 10^{-3}$ | $2.59 \times 10^{-3}$ ($1.35 \times 10^{-3}$–$3.50 \times 10^{-3}$) | 0.12 | 0.121 (0.114–0.131) | 0.017 |
| D | 1 | $3.00 \times 10^{-3}$ | $3.18 \times 10^{-3}$ ($1.24 \times 10^{-3}$–$5.59 \times 10^{-3}$) | 0.20 | 0.197 (0.191–0.207) | 0.003 |
| D | 2 | $7.00 \times 10^{-3}$ | $6.24 \times 10^{-3}$ ($5.17 \times 10^{-3}$–$6.86 \times 10^{-3}$) | 0.04 | 0.043 (0.040–0.049) | 0.003 |
| D | 3 | $3.50 \times 10^{-3}$ | $2.86 \times 10^{-3}$ ($1.85 \times 10^{-3}$–$4.04 \times 10^{-3}$) | 0.16 | 0.159 (0.151–0.167) | 0.003 |
| D | 4 | $8.00 \times 10^{-4}$ | $5.72 \times 10^{-4}$ ($4.16 \times 10^{-4}$–$9.23 \times 10^{-4}$) | 0.06 | 0.061 (0.058–0.065) | 0.003 |
| D | 5 | $2.50 \times 10^{-3}$ | $2.01 \times 10^{-3}$ ($1.37 \times 10^{-3}$–$2.89 \times 10^{-3}$) | 0.13 | 0.131 (0.122–0.137) | 0.003 |

**Table S6. Five-year average transmission-rate and importation-probability estimates. Values summarize the 100 chain-level five-year averages.**

| | | $\beta_g$ | | | $C_{0,g}$ | | |
|---|---|---|---|---|---|---|---|
| Organism | Block | Median | 95% interval | Interquartile range | Median | 95% interval | Interquartile range |
| MRSA | 1 | $1.41 \times 10^{-2}$ | $9.63 \times 10^{-3}$–$1.91 \times 10^{-2}$ | $1.17 \times 10^{-2}$–$1.63 \times 10^{-2}$ | 0.171 | 0.147–0.195 | 0.162–0.180 |
| MRSA | 2 | $8.89 \times 10^{-3}$ | $7.60 \times 10^{-3}$–$1.03 \times 10^{-2}$ | $8.49 \times 10^{-3}$–$9.40 \times 10^{-3}$ | 0.099 | 0.081–0.120 | 0.092–0.103 |
| MRSA | 3 | $8.87 \times 10^{-3}$ | $7.08 \times 10^{-3}$–$1.05 \times 10^{-2}$ | $8.41 \times 10^{-3}$–$9.68 \times 10^{-3}$ | 0.082 | 0.062–0.102 | 0.075–0.088 |
| MRSA | 4 | $2.86 \times 10^{-3}$ | $2.02 \times 10^{-3}$–$3.69 \times 10^{-3}$ | $2.55 \times 10^{-3}$–$3.25 \times 10^{-3}$ | 0.170 | 0.150–0.195 | 0.163–0.179 |
| MRSA | 5 | $7.84 \times 10^{-3}$ | $4.67 \times 10^{-3}$–$9.82 \times 10^{-3}$ | $6.98 \times 10^{-3}$–$8.66 \times 10^{-3}$ | 0.113 | 0.093–0.140 | 0.105–0.120 |
| LRPA | 1 | $1.63 \times 10^{-2}$ | $1.00 \times 10^{-2}$–$2.39 \times 10^{-2}$ | $1.39 \times 10^{-2}$–$1.92 \times 10^{-2}$ | 0.150 | 0.135–0.161 | 0.145–0.156 |
| LRPA | 2 | $8.87 \times 10^{-3}$ | $7.61 \times 10^{-3}$–$1.02 \times 10^{-2}$ | $8.37 \times 10^{-3}$–$9.48 \times 10^{-3}$ | 0.120 | 0.102–0.135 | 0.114–0.125 |
| LRPA | 3 | $1.50 \times 10^{-3}$ | $6.97 \times 10^{-4}$–$2.36 \times 10^{-3}$ | $1.24 \times 10^{-3}$–$1.78 \times 10^{-3}$ | 0.086 | 0.076–0.098 | 0.083–0.090 |
| LRPA | 4 | $4.93 \times 10^{-3}$ | $3.51 \times 10^{-3}$–$7.23 \times 10^{-3}$ | $4.38 \times 10^{-3}$–$5.57 \times 10^{-3}$ | 0.226 | 0.186–0.259 | 0.218–0.237 |
| LRPA | 5 | $9.59 \times 10^{-3}$ | $7.90 \times 10^{-3}$–$1.12 \times 10^{-2}$ | $8.82 \times 10^{-3}$–$1.01 \times 10^{-2}$ | 0.119 | 0.105–0.136 | 0.114–0.124 |
| CRKP | 1 | $5.31 \times 10^{-3}$ | $1.40 \times 10^{-3}$–$7.27 \times 10^{-3}$ | $4.38 \times 10^{-3}$–$6.10 \times 10^{-3}$ | 0.058 | 0.052–0.069 | 0.056–0.060 |
| CRKP | 2 | $2.32 \times 10^{-3}$ | $1.29 \times 10^{-3}$–$3.80 \times 10^{-3}$ | $1.99 \times 10^{-3}$–$2.88 \times 10^{-3}$ | 0.013 | 0.010–0.016 | 0.012–0.014 |
| CRKP | 3 | $2.05 \times 10^{-3}$ | $7.90 \times 10^{-4}$–$3.14 \times 10^{-3}$ | $1.50 \times 10^{-3}$–$2.33 \times 10^{-3}$ | 0.050 | 0.044–0.057 | 0.047–0.053 |
| CRKP | 4 | $2.10 \times 10^{-3}$ | $1.36 \times 10^{-3}$–$2.86 \times 10^{-3}$ | $1.87 \times 10^{-3}$–$2.33 \times 10^{-3}$ | 0.062 | 0.051–0.075 | 0.058–0.066 |
| CRKP | 5 | $2.47 \times 10^{-3}$ | $1.06 \times 10^{-3}$–$4.62 \times 10^{-3}$ | $1.76 \times 10^{-3}$–$3.17 \times 10^{-3}$ | 0.039 | 0.029–0.049 | 0.034–0.043 |
| VRE | 1 | $8.95 \times 10^{-3}$ | $7.93 \times 10^{-4}$–$2.02 \times 10^{-2}$ | $3.57 \times 10^{-3}$–$1.29 \times 10^{-2}$ | 0.412 | 0.340–0.462 | 0.380–0.442 |
| VRE | 2 | $2.70 \times 10^{-3}$ | $6.05 \times 10^{-4}$–$5.22 \times 10^{-3}$ | $1.94 \times 10^{-3}$–$3.80 \times 10^{-3}$ | 0.058 | 0.041–0.073 | 0.053–0.064 |
| VRE | 3 | $4.91 \times 10^{-3}$ | $1.96 \times 10^{-3}$–$6.55 \times 10^{-3}$ | $4.04 \times 10^{-3}$–$5.48 \times 10^{-3}$ | 0.136 | 0.121–0.165 | 0.130–0.142 |
| VRE | 4 | $6.28 \times 10^{-3}$ | $3.46 \times 10^{-3}$–$8.36 \times 10^{-3}$ | $5.30 \times 10^{-3}$–$7.12 \times 10^{-3}$ | 0.183 | 0.141–0.246 | 0.164–0.202 |
| VRE | 5 | $7.48 \times 10^{-3}$ | $5.18 \times 10^{-3}$–$9.72 \times 10^{-3}$ | $6.77 \times 10^{-3}$–$8.24 \times 10^{-3}$ | 0.083 | 0.068–0.100 | 0.078–0.090 |

**Table S7. Empirical coverage of observed weekly detection counts by the 95% fitted-model simulation interval. Each time series contains 261 weeks. The interval for each organism-week was calculated from 50,000 observation draws: 500 Poisson draws from each of 100 fitted-chain latent simulations.**

| Organism | Weeks within interval | Weeks outside interval | Total weeks | Coverage |
|---|---|---|---|---|
| MRSA | 258 | 3 | 261 | 98.9% |
| LRPA | 254 | 7 | 261 | 97.3% |
| CRKP | 258 | 3 | 261 | 98.9% |
| VRE | 251 | 10 | 261 | 96.2% |

**Table S8. Block-specific importation and transmission contributions used for Fig. 5. Contributions are expected colonized patient-weeks from 2012–2016 per 1,000 active patient-weeks in the corresponding block. Percent gives each block's share of total five-year colonization burden for that organism.**

| Organism | Block | Importation | Transmission | Share of total burden |
|---|---|---|---|---|
| MRSA | 1 | 144.9 | 28.3 | 48.2% |
| MRSA | 2 | 81.2 | 44.5 | 15.7% |
| MRSA | 3 | 64.3 | 62.6 | 4.9% |
| MRSA | 4 | 120.3 | 32.5 | 26.9% |
| MRSA | 5 | 71.4 | 42.3 | 4.2% |
| LRPA | 1 | 94.1 | 24.5 | 41.2% |
| LRPA | 2 | 81.8 | 30.2 | 17.5% |
| LRPA | 3 | 51.2 | 8.0 | 2.9% |
| LRPA | 4 | 113.4 | 42.0 | 34.1% |
| LRPA | 5 | 60.5 | 33.3 | 4.3% |
| CRKP | 1 | 45.4 | 7.0 | 49.8% |
| CRKP | 2 | 10.8 | 2.4 | 5.7% |
| CRKP | 3 | 40.4 | 6.2 | 6.2% |
| CRKP | 4 | 43.9 | 13.0 | 34.2% |
| CRKP | 5 | 26.5 | 6.4 | 4.1% |
| VRE | 1 | 253.0 | 23.0 | 68.1% |
| VRE | 2 | 37.8 | 5.5 | 4.8% |
| VRE | 3 | 81.8 | 22.6 | 3.6% |
| VRE | 4 | 87.9 | 49.6 | 21.4% |
| VRE | 5 | 39.9 | 23.9 | 2.1% |
